# Insights from nine nights of self-applied, low-density sleep EEG during sleep restriction therapy for insomnia: a proof-of-concept evaluation

**DOI:** 10.64898/2026.05.08.26348885

**Authors:** Emily C. Stanyer, Maëlwenn Le Roux, Rachel Sharman, Sofia I. Ribeiro Pereira, Shaun Davidson, Lionel Tarassenko, Colin A. Espie, Simon D. Kyle

## Abstract

**Objectives:** Self-applied, low-density EEG offers opportunities to examine sleep in the home environment, yet its feasibility during behavioural sleep interventions remains unexplored. This pilot study aimed to evaluate the feasibility and acceptability of a self-applied, low-density EEG device during sleep restriction therapy (SRT) and explore effects on sleep and affect.

**Methods:** Seventeen adults with insomnia and depressive symptoms completed a 2-week baseline and 4 weeks of SRT. The primary outcome was the proportion of expected EEG recordings completed and scoreable. Secondary outcomes included clinical measures, sleep continuity (sleep diary, actigraphy), sleep architecture (low-density EEG for 9 nights), power spectral density, and affect. Data were analysed with linear mixed models. Cohen’s d and 95% confidence intervals were reported.

**Results:** Feasibility was demonstrated (92% of expected EEG nights completed). SRT was associated with reductions in insomnia severity, depressive symptoms, negative affect, and increases in positive affect. Robust improvements were observed across treatment in sleep continuity (SOL, WASO, SE) from diary, which were paralleled by actigraphy. EEG revealed reduced TIB, TST, N1, N2, REM sleep, and REM latency during week one. Reductions in EEG-derived TIB and N1 sleep were maintained at night 28. There were no reliable differences for spectral or spindle measures.

**Conclusions:** These findings suggest that self-applied, low-density EEG during SRT is feasible, acceptable, and may capture sleep changes during treatment. They highlight the potential for multi-night monitoring of sleep interventions at home and elucidating mechanisms underlying therapeutic change.

**Statement of Significance:** Insomnia, frequently co-occurring with depression, is effectively treated in many patients with cognitive behavioural therapy for insomnia (CBT-I), yet treatment mechanisms are poorly understood, and a large minority do not show clinical improvement. Sleep restriction therapy, a core component of CBT-I, shows promise for improving both sleep and depressive symptoms. This study demonstrates that at-home, self-applied, low-density sleep EEG monitoring during sleep restriction therapy is both practical and well accepted by participants, capturing early changes in sleep outside the laboratory. This approach provides a novel opportunity to monitor treatment effects in real-world settings, tailor interventions, and investigate mechanisms linking sleep and mental health, highlighting the translational potential of wearable sleep technology for advancing both research and clinical care.

## 1. Introduction

Insomnia is a highly prevalent sleep disorder which significantly impairs daytime functioning and quality of life^1^. It frequently co-occurs with depression, contributing to both the onset and persistence of depressive symptoms^2^. While cognitive behavioural therapy for insomnia (CBT-I) is effective in treating insomnia symptoms for many, around 30% of patients remain non-responders^3^. Further understanding and monitoring of the treatment process is required. For example, such monitoring could help distinguish between a failure to engage proposed therapeutic mechanisms and a lack of treatment suitability.

Sleep restriction therapy (SRT), a core component of CBT-I, aims to improve sleep by regularising and limiting time in bed (TIB) to match self-reported sleep duration^4^. SRT has been shown to improve sleep quality, reduce insomnia severity, and be a cost-effective intervention^5–7^. Emerging evidence further suggests that SRT may alleviate depressive symptoms and reduce negative affect, supporting its potential as a transdiagnostic treatment for co-morbid insomnia and depression^8–10^. However, the mechanisms through which SRT exerts its effects on sleep and mood remain poorly understood^11^. Proposed pathways include increased homeostatic sleep pressure, circadian realignment, reductions in physiological arousal, and decreased sleep-related anxiety^11–14^. A greater understanding of these processes is critical for refining SRT protocols, improving treatment outcomes, and identifying which patients are most likely to benefit^15^.

Most previous research relies on broad pre-post comparisons, which can overlook the acute, night-to-night changes in sleep that might occur during early treatment^16–18^. Capturing these nightly changes may provide more precise insight into the mechanisms of SRT, particularly during the initial week. This phase represents a critical window for investigation. It is characterised by intensive TIB restriction and potential side-effects, particularly increased sleepiness and reduced psychomotor vigilance, which may create barriers to adherence^19^.

Advances in low-density, ambulatory electroencephalography (EEG) devices offer a unique opportunity to examine sleep in the home environment. Unlike traditional polysomnography (PSG), these devices are often minimally invasive with few electrodes, allow repeated nightly recordings, and can be self-applied by the individual^20,21^. This enables ecologically valid assessments whilst reducing laboratory confounds, such as the first-night effect^22^. These devices can also capture power spectral density (PSD) measures, providing a window into sleep microstructure. For example, quantifying power within the slow wave activity (SWA) range serves as a proxy for homeostatic sleep pressure, allowing tracking of accumulation and dissipation of sleep debt in response to treatment.

To the best of our knowledge, no study has used ambulatory, self-applied low-density EEG to track sleep changes during the early stages of behavioural sleep treatment. The present pilot study addresses this gap by evaluating the feasibility and acceptability of home EEG recordings in individuals with insomnia disorder and depressive symptoms undergoing SRT. Secondary aims include examining nightly, early change in sleep architecture, power spectral EEG, sleep spindles, and weekly changes in positive and negative affect, providing a more detailed characterisation of the acute processes associated with treatment implementation. By advancing the utility of home-based sleep monitoring methods, this work paves the way for future work to deepen mechanistic understanding of how SRT impacts both sleep and mental health.

## 2. Methods

### 2.1. Study Design

The RESTORE (Sleep RESTriction therapy fOR insomnia in people with dEpressive symptoms) study was a within-subjects, pilot, feasibility study. This study was conducted in Oxfordshire, UK and was approved by the Medical Sciences Interdivisional Research Ethics Committee at the University of Oxford (Reference: R91701/RE001). Informed consent was obtained from all participants. The study was pre-registered on Open Science Framework (https://osf.io/3meuz).

### 2.2. Participants

According to CONSORT guidelines for pilot and feasibility trials, sample sizes should be based on the number of participants required to evaluate study procedures rather than power calculations^23^. Following this, 15 participants were deemed sufficient.

Participants were recruited between April and October 2024 in Oxfordshire (UK), from the community through advertisement (social media, public noticeboards, university websites, and mailing lists). The inclusion criteria were as follows: aged 18-65 years, meeting the DSM-5 criteria for chronic insomnia disorder as assessed by a Sleep Condition Indicator (SCI) score of ≤16 and semi-structured screening interview. Participants had to score ≥8 on the Hospital Anxiety and Depression Scale (depression sub-scale; HADS-D;^24^). Participants were required to typically sleep between the hours of 22:00 and 08:00, and have a self-reported sleep efficiency (SE) of ≤ 85% measured using items 1,3, and 4 from the Pittsburgh Sleep Quality Index ^25^.

Exclusion criteria included: current alcohol misuse (assessed by the Alcohol Use Disorders Identification Test; AUDIT^26^); sleep disorders other than insomnia disorder (assessed by bespoke sleep disorders questions based on Klingman et al.^27^); current shift work; prescribed medication for insomnia, mood, or any that (in the opinion of the investigator) could affect sleep; medical comorbidity (chronic migraine, epilepsy); contraindication to wet-gel ECG electrodes’ a pacemaker (due to use of Bluetooth within the EEG device); current psychotherapy for sleep/insomnia; international travel across 2 or more time zones during the study; previous experience of SRT; pregnancy, or planning pregnancy in next two months; current suicidal ideation with intent or recent attempt (> mild risk on the Columbia Suicide Severity Rating Scale; C-SSRS;^28^); recreational drug use in the past three months; and residence outside the designated study area. Participants were reimbursed £150 in the form of Amazon vouchers.

### 2.3. Procedures

See Figure 1 for the study timeline. Individuals interested in the study completed an online screening questionnaire (Qualtrics^XM^, Provo, UT), followed by a semi-structured telephone interview to determine eligibility. Eligible participants attended a baseline visit, during which they completed questionnaires and were introduced to study equipment. Participants were provided with an actigraphy device (MotionWatch8, CamNtech) to wear continuously for six weeks on their non-dominant wrist. Participants completed a digital morning and evening sleep diary alongside the Positive and Negative Affect Schedule (PANAS) twice daily. Participants self-applied the low-density EEG device for two consecutive nights (adaptation night, baseline night) at the end of the baseline period, seven consecutive nights during the first treatment week, and one night at the end of treatment (night 28). Participants were instructed to apply the device up to 15 minutes before habitual bedtime. At the end of treatment, participants also completed follow-up questionnaires.

**Figure 1:**
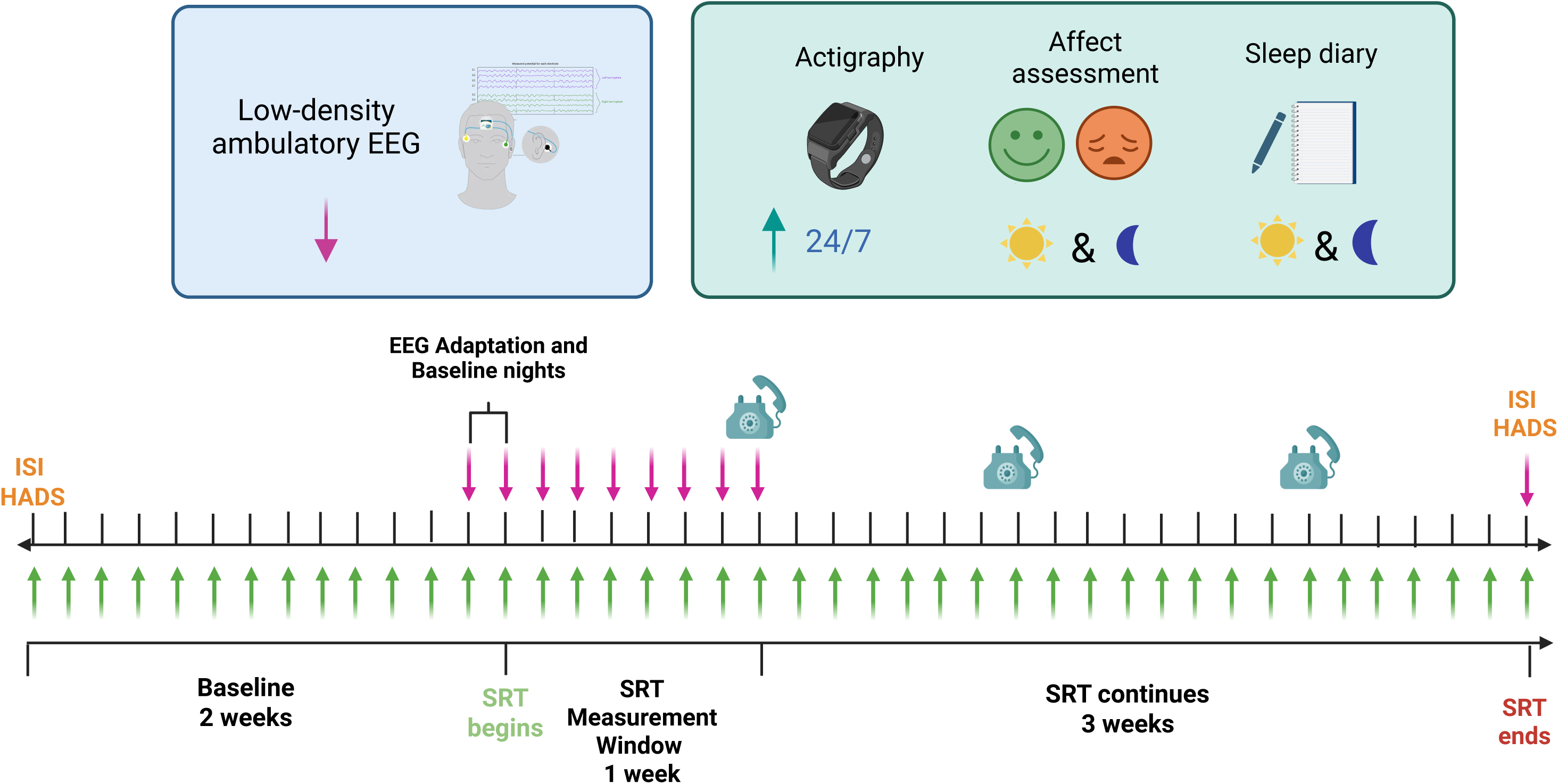
RESTORE Study timeline. Please note that a modified version of this figure is published elsewhere^58^.

### 2.4. Sleep Restriction Therapy

Sleep restriction therapy was delivered 1:1 in a one-hour session by a researcher (E.C.S.) trained in CBT-I under the supervision of experts in sleep medicine (S.D.K., C.A.E.). Sessions were guided with a standardised protocol including workbooks, personalised treatment slides, and sleep hygiene information according to previous protocols^6^. This included advice on maximising morning/daytime light exposure, minimising evening light exposure, reducing stimulant use such as nicotine and caffeine. Participants were advised not to nap; however, if it was necessary, they were told to do so for only 20-30 minutes in the early afternoon (13:00-15:00). The prescribed TIB was taken from average self-reported total sleep time (TST) calculated from the participant’s baseline sleep diary. The minimum possible prescribed TIB was 5 hours. Risetime was determined in discussion with the researcher based on a combination of typical risetime, personal preference, and childcare/work commitments. Subsequently, the bedtime was set in accordance with the prescribed TIB. Participants received four, weekly 15-minute phone calls to adjust sleep windows based on the self-reported SE from the sleep diary from the previous week. Adjustments to TIB were made as follows:

- SE ≥85%: Increase TIB by 15 minutes.
- SE <80%: Decrease TIB by 15 minutes.
- SE 80-84%: Maintain current sleep schedule.

Consistent with clinical protocols^6^, TIB prescription was adapted (increased) in collaboration with the participant if they experienced side-effects or substantial difficulty with adherence. Participants received four guided weeks of SRT but were encouraged to continue after the end of the formal treatment phase in line with guidelines^29^.

### 2.5. Outcome Measures

Study data were collected and managed using Research Electronic Data Capture (REDCap) electronic data capture tools hosted at the University of Oxford.

#### 2.5.1. Primary Outcomes

The primary outcomes were: (1) the proportion of expected EEG recordings that were completed and, among test nights (Baseline, Nights 1-7 SRT, Night 28), the number that yielded scorable, usable data; and (2) participant acceptability of the low-density EEG. For scoreability, each EEG recording was rated by the researcher on a 0-10 scale. A score of 0 indicated a non-scorable recording, typically characterised by persistent high-amplitude artefacts (e.g., gross body movement), sustained high electrode impedances, or signal loss that obscured the EEG/EMG. Conversely, a score of 10 indicated a highly scoreable recording with a clear signal-to-noise ratio, allowing for the easy identification of sleep features (eye movements, K-complexes, spindles) and stages. The number of EEG nights completed by participants was reported for the first week and across the entire study period (adaptation, baseline, nights 1-7, and night 28). Thus, the mean number of completed EEG nights across participants is reported out of 10 expected nights. In terms of acceptability, at follow-up (day 29), participants completed a telephone feedback interview comprising a bespoke question rated on a 0-10 scale: “Please rate your discomfort while wearing the EEG head device (0 = not uncomfortable at all, 10 = extremely uncomfortable).”

#### 2.5.2. Secondary Outcomes

##### 2.5.2.1. Treatment Adherence and Adverse Events

Adherence to the prescribed sleep schedule was assessed by comparing prescribed bed and rise times to those recorded in the sleep diary and derived from actigraphy. The number of returned, incomplete, or missing diaries was recorded, with incomplete diaries defined as having <50% of days with both bedtime and risetime reported. Participants were considered adherent if the self-reported or actigraphy-derived bedtime was no more than 15 minutes earlier than prescribed, and if the self-reported or actigraphy-derived risetime was no more than 15 minutes later than the prescribed risetime. Further details of adherence calculations are provided in the supplement. Although not pre-specified, the number, duration, and start time of self-reported naps was summarised for both the baseline and treatment period.

Serious Adverse Events (SAEs) were defined as any untoward medical occurrence that resulted in:

- Death or life-threatening circumstances;
- Inpatient hospitalisation or prolongation of existing hospitalisation;
- Persistent or significant disability or incapacity;
- A congenital anomaly or birth defect.

Adverse events presumed to be related to the EEG device were recorded.

##### 2.5.2.2. Baseline Questionnaires

Chronotype was assessed using the full 19-item Morningness-Eveningness Questionnaire (MEQ^30^), and caffeine intake via the Caffeine Consumption Questionnaire-Revised (CCQ-R^31^).

##### 2.5.2.3. Insomnia Severity and Depressive Symptoms

Insomnia severity was assessed pre and post-treatment using the Insomnia Severity Index (ISI^32^; α = 0.83^32^), and depressive symptoms using the HADS depression subscale (HADS-D; α = 0.84^33^), with scores categorized as <8 = none, 8-10 = mild, 11-15 = moderate, >15 = severe.

##### 2.5.2.4. Positive and Negative Affect

Positive and negative affect were measured using the Positive and Negative Affect Schedule (PANAS^34^; α = 0.89/0.85^35^), completed once at baseline and twice daily in morning and evening diaries throughout the study. The timing of PANAS completion was not constrained, allowing for retrospective entry, similar to a paper survey. To evaluate completion timings, the intervals between morning survey completion and self-reported rise time, as well as between evening survey completion and self-reported bedtime, were calculated. These were reported as mean hours and minutes (HH:MM) and standard deviation (SD).

##### 2.5.2.5. Sleep Diary and Actigraphy-derived Sleep Continuity

Participants completed an online sleep diary based on the Consensus Sleep Diary^36^ in the morning after waking up and in the evening before bedtime for two weeks during baseline, and for four weeks during treatment. This was completed on the same survey as the PANAS. Participants could report the number of naps, the duration and timing of these, as well as any periods in which they took their actiwatch off. Measures derived from the sleep diary included: TIB, TST, wake time after sleep onset (WASO; minutes), sleep onset latency (SOL; minutes), SE (%). Actigraphy data were scored using MotionWare Software (v1.3.17; CamNtech Ltd.) following a pre-specified analysis protocol. A description of the scoring procedure is provided in the supplement. Actigraphy measures included TIB, TST, WASO, SOL, SE, and sleep midpoint (HH:MM).

##### 2.5.2.6. EEG-derived Sleep Architecture

Participants self-applied a low-density ambulatory EEG device (HomeSleepTest REM+, SOMNOmedics). The device included a single frontopolar EEG channel (Approximately Fpz-M1), two electro-oculogram (EOG) channels, and one mastoid electromyogram (EMG; M1). The device also recorded movement, head position, ambient light, and continuous impedance. Data were recorded at a sampling rate of 256 Hz and filtered between 0.3 and 35 Hz in accordance with American Academy of Sleep Medicine (AASM) guidelines Version 3.0^37^.

Participants wore the device for an adaptation night and a baseline night, during which they were instructed to maintain their habitual sleep-wake schedule. During the first week of SRT, participants wore the device for seven consecutive nights, beginning on the night of the initial SRT session, and then for one further night at the end of the treatment period. The adaptation night was not scored and thus not included in the primary outcome of scoreability nor the secondary outcomes analyses.

Participants uploaded their recordings to an encrypted cloud server managed by SOMNOmedics, from which data were subsequently downloaded for analysis. Scoring adhered to the AASM guidelines Version 3.0^37^ where possible and was conducted in DOMINO™ in 30 second epochs (SOMNOmedics; Version 3.0.0.8). Artefact epochs were visually identified and the mean global artefact percentage across all recordings was reported. Sleep architecture measures included: TIB, TST, SOL, WASO, SE, the absolute duration of N1, N2, N3, REM sleep, N3 sleep latency, and REM sleep latency (all minutes). Recordings were scored by an experienced sleep scorer (E.C.S.) blinded to timepoint. The researcher was trained on a random, blinded subset (10%) of recordings by an experienced, accredited somnologist (R.S.).

##### 2.5.2.7. Power Spectral Analysis

A power spectral analysis was performed on the single EEG channel based on N2 and N3 sleep epochs free of visually identified EEG and EMG artefacts or movement arousals. As the number of epochs differed between baseline and treatment nights, we calculated the maximal number of artefact-free NREM sleep epochs common to all nights and participants, and hypnograms were trimmed to the resulting common maximum as in previous studies^38,39^. Analysis of theta band activity was performed during REM epochs, on the maximum common number of REM epochs. Contrary to the pre-registration, PSD analysis was conducted using custom-made MATLAB^©^ scripts using Fieldtrip^40^ in MATLAB^©^ 2025, rather than the SpiSOP toolbox to allow greater customisation. For computing individual average power spectra, the data from NREM sleep epochs were divided into consecutive 4-s segments with a 50% overlap. Using Fieldtrip’s *mtmfft* function, each segment was tapered using a sequence of Slepian windows before the application of a Fast Fourier Transform that resulted in segment power spectra with a frequency resolution of 0.25 Hz. Power spectra were then averaged across all segments (Welch’s method) to obtain Power Spectral Density (PSD) estimates. Band power was computed by integrating the PSD using Simpson’s rule across the following frequency bands of interest: slow-wave activity (SWA, 0.5-4.5Hz), delta (0.75-4.5Hz), theta (4.5-8.0Hz), alpha (8.0-12.0Hz), sigma (12.0-15Hz), and beta (15.0-32.0Hz). Normalised percentage band power was then computed by dividing the power in each band by the PSD integrated across the entire range of frequencies of interest (0.5 Hz-32.0 Hz).

Sleep spindles and slow oscillations (SO) were detected from the Fpz channel during the maximal common number of artefact-free NREM epochs using the SpiSOP MATLAB^©^ toolbox (https://www.spisop.org).

### Spindle Detection

Although visual identification of spindles was pre-specified, spindle detection in the current version of SpiSOP is performed automatically. The identified spectral peak was below the expected spindle range (likely due to the frontal derivation) and did not differentiate between fast and slow spindles. Therefore, spindle detection was conducted using a standardised centre frequency of 12 Hz). A sliding window (0.2 s) was used to calculate the root mean square (RMS) of the band-pass filtered signal with detections defined by a smoothed RMS exceeding 1.5 SD of the filtered signal. Spindle duration was restricted to 0.5-3 s, with an amplitude ceiling of 200 µV. Total spindle count and spindle density were calculated; the latter defined as the mean number of spindles per 30-second NREM epoch.

### Slow Oscillation Detection

The signal was further band-pass filtered (0.3-3.5 Hz) to identify consecutive zero-crossings corresponding to a waveform in the 0.5-1.11 Hz frequency range. SO candidates were excluded if trough amplitude was greater than −15 µV (i.e., less negative than −15 µV), positive peak amplitude was < 10 µV, or peak-to-peak amplitude exceeded 1000 µV. Final selection required a trough and total amplitude >1.25 times the mean of all putative detections. Total SO count and SO density were calculated, the latter defined as the mean number of SOs per 30-second NREM epoch.

#### 2.5.3. Statistical Analysis

CONSORT guidelines^23^ do not recommend hypothesis testing for pilot studies on the basis that they are underpowered to detect differences. Thus, we focus on descriptive reporting (mean differences, 95% confidence intervals (CIs), and effect sizes (Cohen’s *d*)). Differences were considered reliable if the 95% CIs did not cross zero. Cohen’s *d* was calculated as the within-subject treatment effect divided by the standard deviation (SD) of the change score from baseline using complete cases. Researchers were blinded to timepoint and participant identifiers during analysis. Statistical analyses were conducted in RStudio (version: 2026.01.0-392) and the scripts are available on GitHub (https://github.com/emstanyer/The-RESTORE-Study).

For the primary outcomes, the completion rate was reported as a percentage (SD) for the overall expected ten nights (including adaptation) and the consecutive seven nights. Scoreability of EEG recordings and participant self-reported discomfort were reported as mean and SD. For secondary outcomes, summary statistics are presented as means and SDs unless otherwise specified. Time values (midpoint of sleep) were analysed as a circular mean. No acceptability or feasibility threshold was determined *a priori*.

Linear mixed models (LMMs) were used to analyse EEG and spectral data, with timepoint (baseline, nights 1-7, night 28) as a fixed effect. A participant-specific random intercept was included to account for repeated measures and their non-independence. LMMs were also conducted on SRT treatment weeks with week as a fixed effect, (baseline, weeks 1-4) for sleep diary, actigraphy, and PANAS. To assess changes in ISI and HADS-D scores between baseline and post-treatment, paired *t*-tests (on complete cases) were performed after verifying that data met the assumptions for parametric testing.

Although it was pre-specified that participants who completed fewer than four consecutive EEG recordings during the first week would be excluded from secondary analyses, all participants were retained for the analysis. Only one participant (two EEG files) would have been excluded had this criterion been applied. Given the preliminary nature of this study, we prioritised maximising data for the assessment of feasibility and descriptive trends.

## 3. Results

### 3.1. Participant Characteristics

Four hundred and ninety-eight individuals completed the online screening. Of these, 26 were eligible to proceed to semi-structured screening interview. Nineteen participants met eligibility criteria, and 17 participants completed the baseline assessments and underwent SRT. All 17 participants completed post-treatment assessments. See Figure 2 for the CONSORT flow diagram.

**Figure 2:**
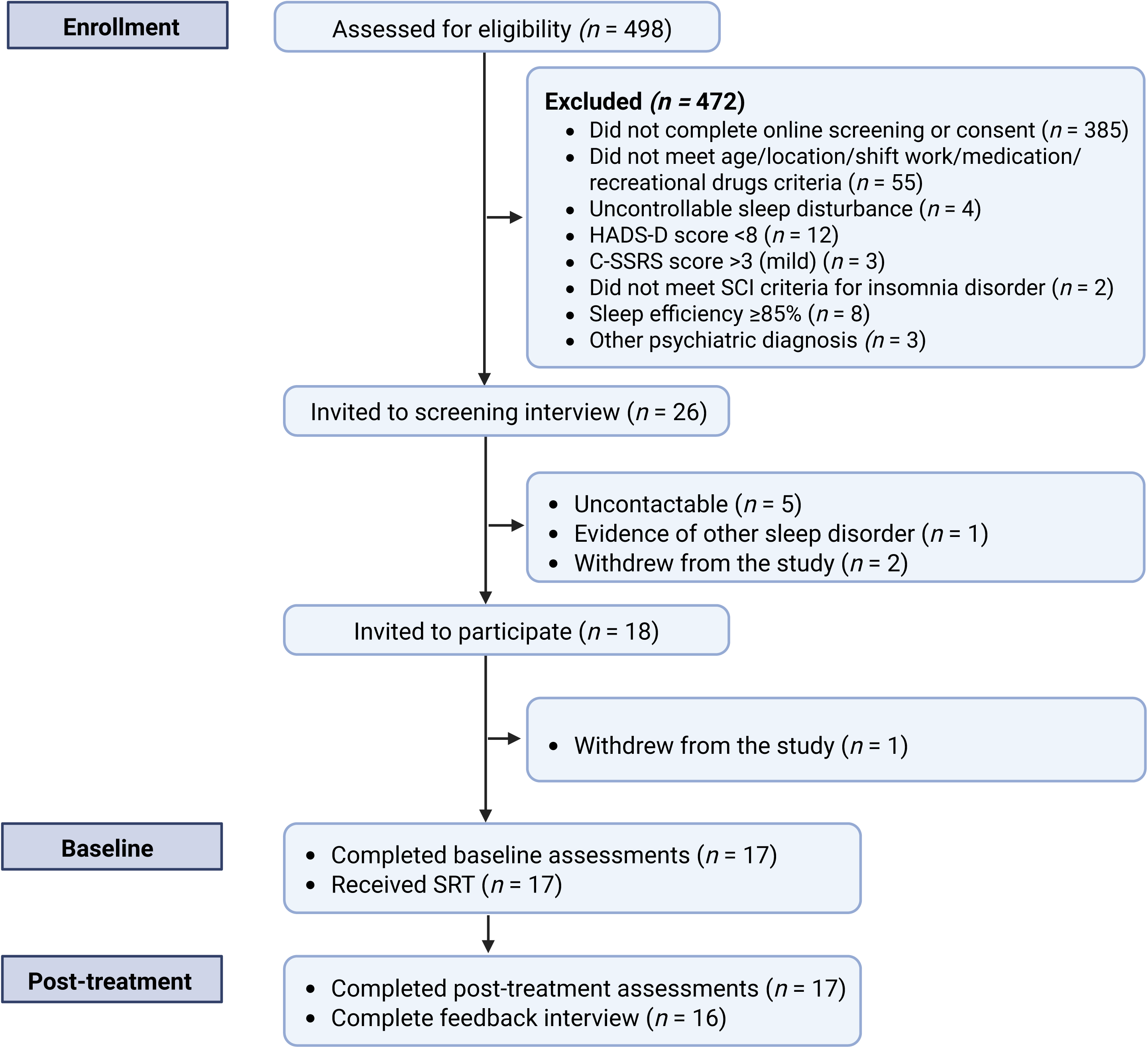
Consolidated Standards of Reporting Trials (CONSORT) flow diagram. SCI = Sleep Condition Indicator, C-SSRS = Columbia Suicide Severity Rating Scale, HADS-D = Hospital Anxiety and Depression Scale Depression Sub-scale, SRT = Sleep restriction therapy.

Baseline sample characteristics are presented in Table 1. The mean age of participants was 38.23 years (SD = 13.89, range = 19-62). Participants were predominantly female (76.47%), university-educated (76.59%), with the majority holding postgraduate qualifications (58.82%). Most were White British (70.64%) and single (52.94%).

**Table 1:** Baseline characteristics.

| Characteristic | Value |
| --- | --- |
| **Number of participants** | 17 |
| **Demographics** |  |
| Age (years), mean (SD) | 38.23 (13.89) |
| Age range | 19 - 62 |
| Male, *n* (%) | 4 (23.53) |
| **Ethnicity, *n* (%)** |  |
| White, British | 12 (70.59) |
| White, Any other white background | 1 (5.88) |
| Asian or Asian British | 0 (0) |
| Black, African, Caribbean, or Black British | 1 (5.88) |
| Mixed or multiple ethnic groups | 2 (11.76) |
| Other ethnic group | 1 (5.88) |
| **Highest level of qualification, *n* (%)** |  |
| None | 0 (0) |
| GCSE or equivalent | 1 (5.88) |
| A-levels or equivalent | 3 (17.65) |
| University undergraduate | 3 (17.64) |
| University postgraduate | 10 (58.82) |
| **Marital status, *n* (%)** |  |
| Single | 9 (52.94) |
| Married or in a domestic partnership | 7 (41.18) |
| Divorced | 1 (5.88) |
| Widowed | 0 (0) |
| Separated | 0 (0) |
| **Depressive symptoms** |  |
| HADS-D score (Mean, SD) | 10.00 (2.76) |
| HADS-D score $\geq 8$ (*n*, %) | 15 (88.24%) |
| **Insomnia severity** |  |
| ISI score (Mean, SD) | 16.77 (3.72) |
| ISI score $\geq 11$ (*n*, %) | 17 (100.00) |
| **Caffeine consumption** |  |
| CCQ-R score (mg/day; Mean, SD) | 184.10 (149.42) |
| CCQ-R score $< 400$mg/day (*n*, %) | 15 (88.24%) |
| CCQ-R score $\geq 400$mg/day (*n*, %) | 2 (11.76%) |
| **Chronotype** |  |
| MEQ score (Mean, SD) | 52.88 (12.73) |
| Morning type (59-86) (*n*, %) | 5 (29.41) |
| Evening type (16-41) (*n*, %) | 3 (17.65) |
| Intermediate type (42-58) (*n*, %) | 9 (52.94) |

Abbreviations: ISI, Insomnia Severity Index, HADS-D, Hospital Anxiety and Depression Scale, (depression sub-scale); CCQ-R, Caffeine Consumption Questionnaire Revised; MEQ, Morning Eveningness Questionnaire; SD, standard deviation.

ISI scores at baseline were within the moderate clinical range (mean = 16.77, SD = 3.72). Sleep diary SE was below 85% (mean = 71.33%, SD = 16.45; Table 2) in line with inclusion criteria. MEQ scores indicated that most participants were intermediate types (*n* [% of sample] 9 [52.94]) %).

**Table 2:** Sleep continuity, insomnia, and depressive symptoms severity.

| **Metric** | **Week** | **Mean (SD)** | **Diff<sub>adj</sub>** | **95% CI** | **d** |
| --- | --- | --- | --- | --- | --- |
| **Sleep Diary** |  |  |  |  |  |
| **(n = 17)** |  |  |  |  |  |
| **TIB (mins)** |  |  |  |  |  |
|  | Baseline | 541.97 (53.73) |  |  |  |
|  | Week 1 | 397.75 (56.31) | -144.22 | -172.86 to -115.58 | -2.52 |
|  | Week 2 | 429.99 (64.30) | -111.97 | -140.61 to -83.33 | -2.50 |
|  | Week 3 | 439.85 (55.27) | -102.12 | -130.76 to -73.48 | -1.71 |
|  | Week 4 | 442.81 (54.12) | -99.16 | -127.80 to -70.52 | -1.76 |
| **TST (mins)** |  |  |  |  |  |
|  | Baseline | 371.04 (61.92) |  |  |  |
|  | Week 1 | 345.11 (54.85) | -25.93 | -47.04 to -4.82 | -0.83 |
|  | Week 2 | 373.35 (50.55) | 2.31 | -18.80 to 23.43 | 0.07 |
|  | Week 3 | 374.01 (50.27) | 2.97 | -18.14 to 24.09 | 0.07 |
|  | Week 4 | 381.95 (54.40) | 10.91 | -10.20 to 32.03 | 0.24 |
| **SOL (mins)** |  |  |  |  |  |
|  | Baseline | 36.05 (24.69) |  |  |  |
|  | Week 1 | 20.59 (17.68) | -15.46 | -24.61 to -6.30 | -0.75 |
|  | Week 2 | 19.47 (23.59) | -16.58 | -25.73 to -7.42 | -0.95 |
|  | Week 3 | 15.81 (15.80) | -20.24 | -29.39 to -11.08 | -1.14 |
|  | Week 4 | 15.61 (15.00) | -20.44 | -29.60 to -11.28 | -1.21 |
| **WASO (mins)** |  |  |  |  |  |
|  | Baseline | 34.99 (26.07) |  |  |  |
|  | Week 1 | 15.75 (11.67) | -19.24 | -32.29 to -6.19 | -0.70 |
|  | Week 2 | 13.13 (11.92) | -21.86 | -34.91 to -8.81 | -1.10 |
|  | Week 3 | 21.45 (26.52) | -13.54 | -26.59 to -0.49 | -0.46 |
|  | Week 4 | 16.90 (17.25) | -18.08 | -31.13 to -5.03 | -0.79 |
| **SE (%)** |  |  |  |  |  |
|  | Baseline | 69.88 (11.59) |  |  |  |
|  | Week 1 | 87.16 (7.06) | 17.28 | 12.19 to 22.36 | 1.62 |
|  | Week 2 | 87.11 (6.94) | 17.22 | 12.14 to 22.31 | 1.73 |
|  | Week 3 | 85.77 (7.04) | 15.89 | 10.80 to 20.98 | 1.37 |
|  | Week 4 | 86.46 (7.33) | 16.58 | 11.49 to 21.66 | 1.37 |
| **Actigraphy** |  |  |  |  |  |
| **(n = 15)** |  |  |  |  |  |
| **TIB (mins)** |  |  |  |  |  |
|  | Baseline | 518.71 (48.58) |  |  |  |
|  | Week 1 | 396.18 (67.82) | -121.60 | -154.09 to -89.12 | -1.85 |
|  | Week 2 | 430.44 (50.85) | -87.34 | -119.82 to -54.86 | -1.59 |
|  | Week 3 | 463.30 (59.55) | -54.49 | -86.97 to -22.00 | -0.72 |
|  | Week 4 | 453.59 (56.94) | -64.19 | -96.68 to -31.71 | -1.03 |
| **TST (mins)** |  |  |  |  |  |
|  | Baseline | 417.35 (43.84) |  |  |  |
|  | Week 1 | 338.84 (50.86) | -78.44 | -104.47 to -52.41 | -1.32 |
|  | Week 2 | 363.50 (36.71) | -53.78 | -79.81 to -27.75 | -1.26 |
|  | Week 3 | 384.98 (40.00) | -32.30 | -58.33 to -6.27 | -0.65 |
|  | Week 4 | 382.19 (42.12) | -35.08 | -61.11 to -9.05 | -0.75 |
| **SOL (mins)** |  |  |  |  |  |
|  | Baseline | 18.39 (15.44) |  |  |  |
|  | Week 1 | 7.08 (4.66) | -11.08 | -18.77 to -3.38 | -0.67 |
|  | Week 2 | 7.85 (10.16) | -10.30 | -18.00 to -2.60 | -0.50 |
|  | Week 3 | 6.29 (5.82) | -11.86 | -19.56 to -4.16 | -0.78 |
|  | Week 4 | 7.57 (6.57) | -10.58 | -18.28 to -2.88 | -0.64 |
| **WASO (mins)** |  |  |  |  |  |
|  | Baseline | 79.64 (21.26) |  |  |  |
|  | Week 1 | 48.51 (19.79) | -30.77 | -41.08 to -20.46 | -3.38 |
|  | Week 2 | 57.11 (21.31) | -22.17 | -32.48 to -11.86 | -2.36 |
|  | Week 3 | 69.67 (33.65) | -9.62 | -19.93 to 0.70 | -0.36 |
|  | Week 4 | 62.42 (25.22) | -16.87 | -27.18 to -6.55 | -1.20 |
| **SE (%)** |  |  |  |  |  |
|  | Baseline | 80.85 (4.72) |  |  |  |
|  | Week 1 | 85.89 (4.19) | 4.93 | 2.79 to 7.07 | 1.61 |
|  | Week 2 | 84.92 (5.49) | 3.95 | 1.81 to 6.09 | 0.89 |
|  | Week 3 | 83.68 (6.23) | 2.72 | 0.58 to 4.86 | 0.48 |
|  | Week 4 | 84.81 (5.18) | 3.85 | 1.71 to 5.99 | 0.97 |
| **Sleep Midpoint** |  |  |  |  |  |
|  | Baseline | 03:04 (01:17) |  |  |  |
|  | Week 1 | 03:14 (01:08) | 00:09 | -00:13 to 00:30 | 0.22 |
|  | Week 2 | 03:12 (01:07) | 00:06 | -00:15 to 00:28 | 0.13 |
|  | Week 3 | 03:03 (01:03) | -00:03 | -00:24 to 00:19 | -0.06 |
|  | Week 4 | 03:04 (01:08) | -00:01 | -00:23 to 00:20 | -0.03 |
| **Insomnia severity (n = 17)** |  |  |  |  |  |
| ISI | Baseline | 16.77 (3.71) |  |  |  |
|  | Week 4 | 7.59 (4.27) | -9.18<sup>†</sup> | -12.10 to -6.27 | -1.62 |
| **Depressive symptoms (n = 17)** |  |  |  |  |  |
| HADS-D | Baseline | 10.00 (2.76) |  |  |  |
|  | Week 4 | 5.59 (4.10) | -4.41<sup>†</sup> | -6.41 to -2.41 | -1.13 |

Abbreviations: Diff<sub>adj</sub>, mean difference adjusted for missing values derived from linear mixed models; 95% CI, 95% confidence interval of the mean difference; *d*, Cohen's *d*; SD, standard deviation; TIB, time in bed; TST, total sleep time; SOL, sleep onset latency; WASO, wake after sleep onset; SE, sleep efficiency; ISI, Insomnia Severity Index, HADS-D, Hospital Anxiety and Depression Scale, (depression sub-scale). <sup>†</sup>The mean difference for paired *t*-tests represents the raw mean difference.

Depression severity scores at baseline were in the clinical range, representing mild depression severity (HADS-D mean = 10.00, SD = 2.76). Mean daily caffeine consumption, assessed using the CCQ-R, was 163 mg/day (SD = 149.42), which is below the recommended daily limit of 400 mg^41^.

### 3.2. Feasibility and Acceptability

The mean completion rate when considering all possible ten EEG recordings across the study was 92.35% (SD = 7.65) with the remaining data being lost due to device failure or participant non-compliance. When considering only the seven consecutive SRT nights the mean completion rate was 89.92% (SD = 10.10). Of the completed total test nights (excluding adaption; expected nights = 153), 16 were deemed not visually scoreable by the researcher. Recording missingness was distributed across nine participants. Reasons for data loss included electrode displacement during the night, high impedances, and excessive movement or artefact obscuring the EEG/EMG signal. The mean researcher-rated scoreability (0-10 scale) of the remaining test recordings (137) was 7.68 out of 10 (SD = 2.27). The average global artefact percentage staged by the researcher across these recordings was 2.68% (SD = 8.02). Regarding acceptability, in the follow-up interview (day 29), participants reported low discomfort when wearing the EEG device, with a mean rating of 2.14 (SD = 1.88; 0 = no discomfort, 10 = extreme discomfort).

### 3.3. Treatment Adherence and Adverse Events

All participants attended every SRT session. Most participants curtailed their TIB through delayed bedtimes and advanced risetimes (Figure S1 A), with TIB gradually increasing across treatment weeks (Table S1). All participants provided sufficient sleep diary data, with none falling below the 50% completeness threshold. Mean sleep diary adherence was 73.87 (SD = 22.44%), higher for bedtimes (85.61% [18.24%]) than risetimes (62.13% [29.48%]), with actigraphy demonstrating a similar pattern (63.88% [20.93%] overall; bedtimes 79.62% [15.74%], risetimes 51.96% [31.63%], Figures S1 C-D). TIB adherence was maintained and generally increased over time (Figure S1 B).

The proportion of participants napping at least once decreased from week 2 of the baseline period (35.3%) and remained stable at 23.5% during SRT weeks 1 and 2. Proportion napping further declined to 11.8% in SRT week 3, with no naps recorded in the final treatment week. Detailed nap characteristics, including start times and durations, are provided in the supplement (Table S2).

No SAEs were reported. Two participants experienced mild skin irritation (e.g., redness, itching) when using the EEG/EMG electrodes. These two instances occurred after five and six consecutive nights of recording, respectively. Following study protocol, both participants were instructed to discontinue use of the EEG device for the remainder of the intervention week. In both cases, symptoms resolved spontaneously within 2-3 days, and both participants successfully completed the final recording night (night 28).

### 3.4. Insomnia Severity and Depressive Symptoms

SRT was associated with reductions in insomnia severity and depressive symptoms. ISI scores decreased from baseline to post-treatment by 9.18 points (*d* = −1.62; Table 2), and HADS-D scores decreased by 4.41 points (*d* = −1.13; Table 2).

### 3.5. Positive and Negative Affect

The mean clock time for completion of the morning PANAS was 08:16 (SD = 03:03) and 22:11 (SD = 03:13) for the evening PANAS. There were increases in morning positive affect in weeks 2 (*d* = 0.34), 3 (*d* = 0.42), and 4 (*d* = 0.52), and decreases in morning negative affect in weeks 3 (*d* = −0.29) and 4 (*d* = −0.27; Table 3; Figure 3). There were increases in evening positive affect in weeks 2 (*d* = 0.44), 3 (*d* = 0.29), and 4 (*d* = 0.41), and a decrease in evening negative affect (*d* = −0.24) in week 3.

**Figure 3:**
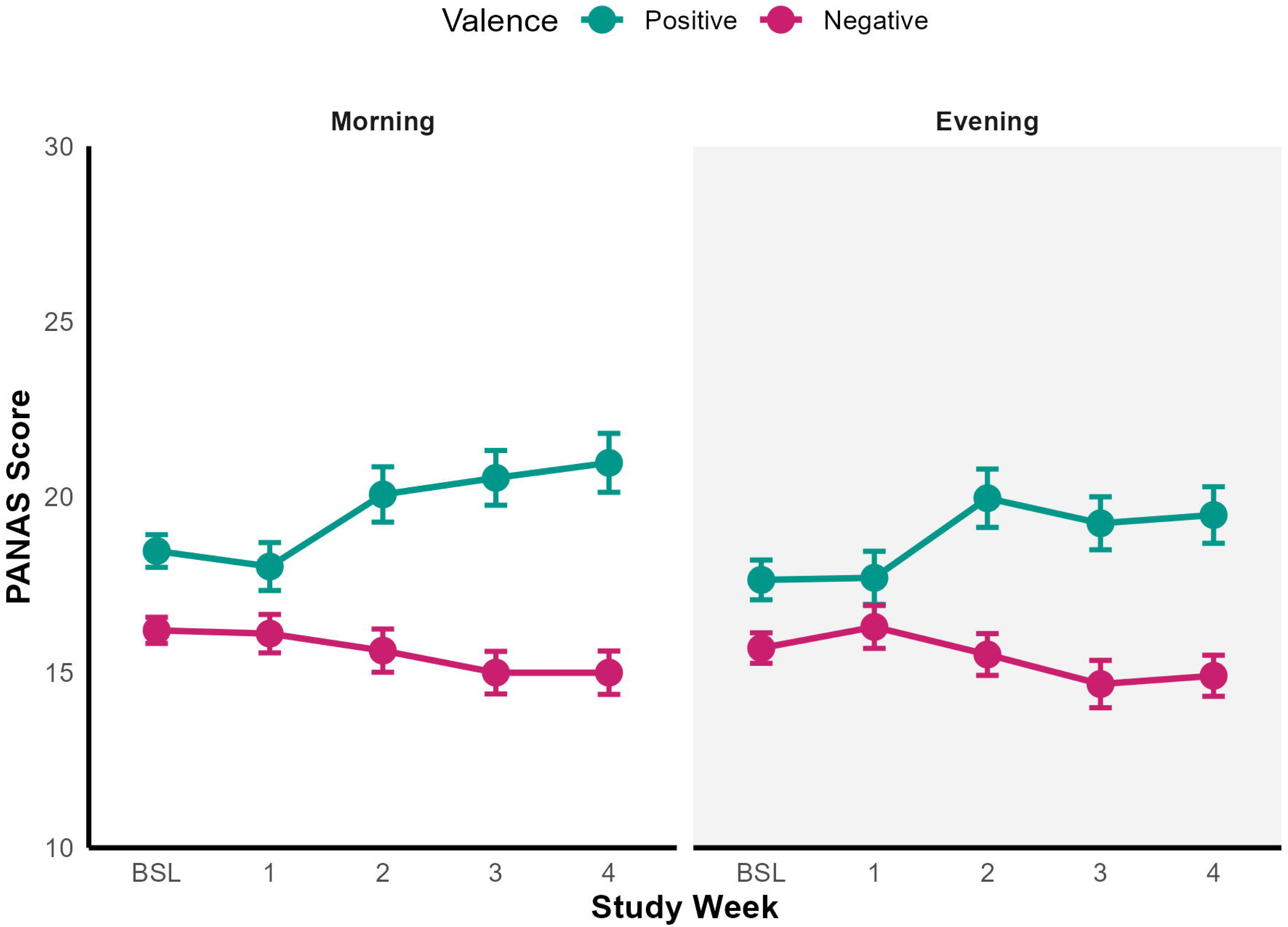
Positive and negative affect (PANAS) taken from morning and evening daily diary pre and throughout SRT. PANAS = Positive and Negative Affect Schedule. Points represent the mean, and error bars represent the standard error (SE).

**Figure 4:**
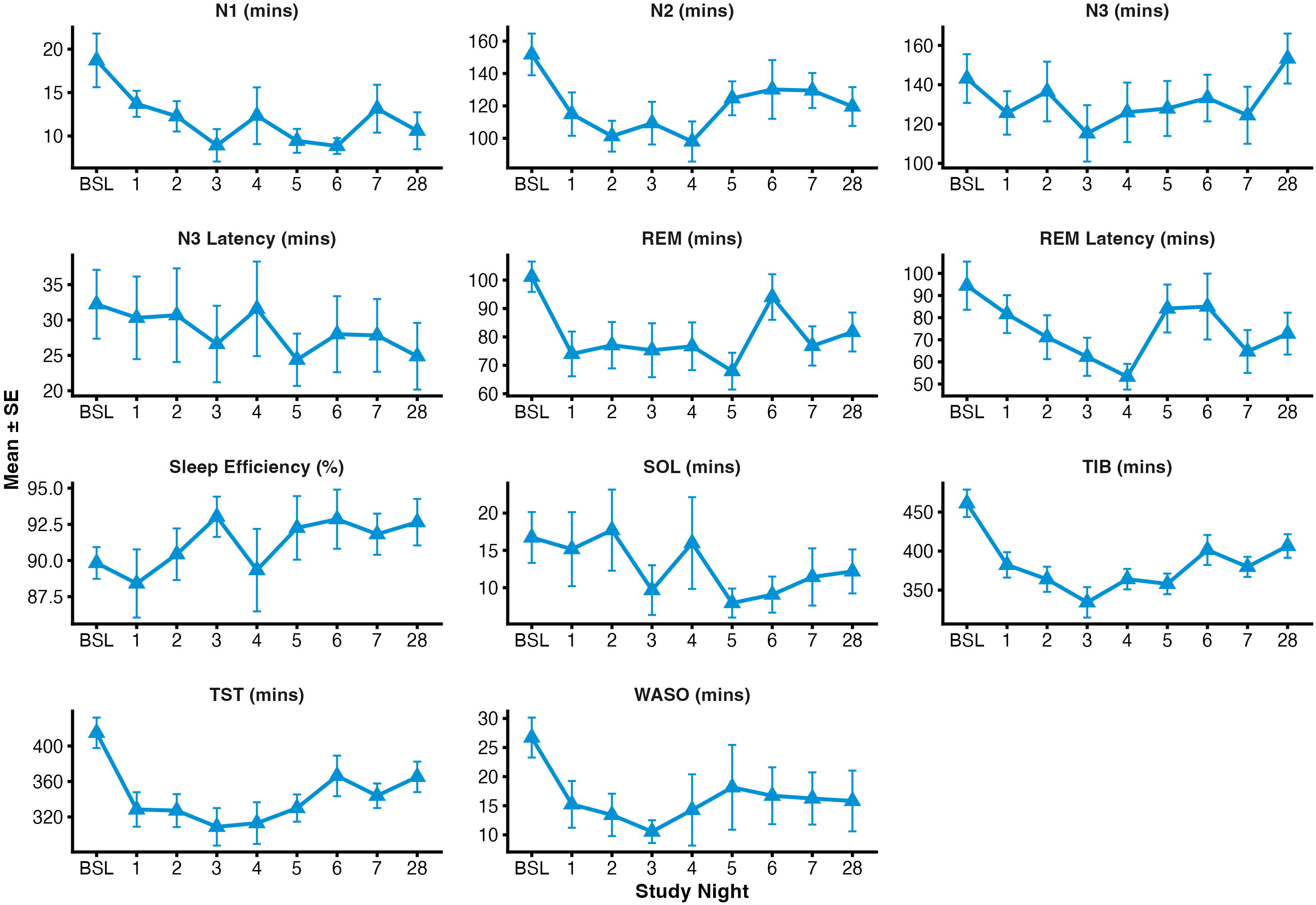
Sleep variables derived from low-density EEG pre, during the first week of SRT, and at night 28. BSL = baseline, TIB = time in bed, WASO = wake after sleep onset, TST = total sleep time, SOL = sleep onset latency. Points represent the mean, and error bars represent the standard error (SE).

**Table 3:**
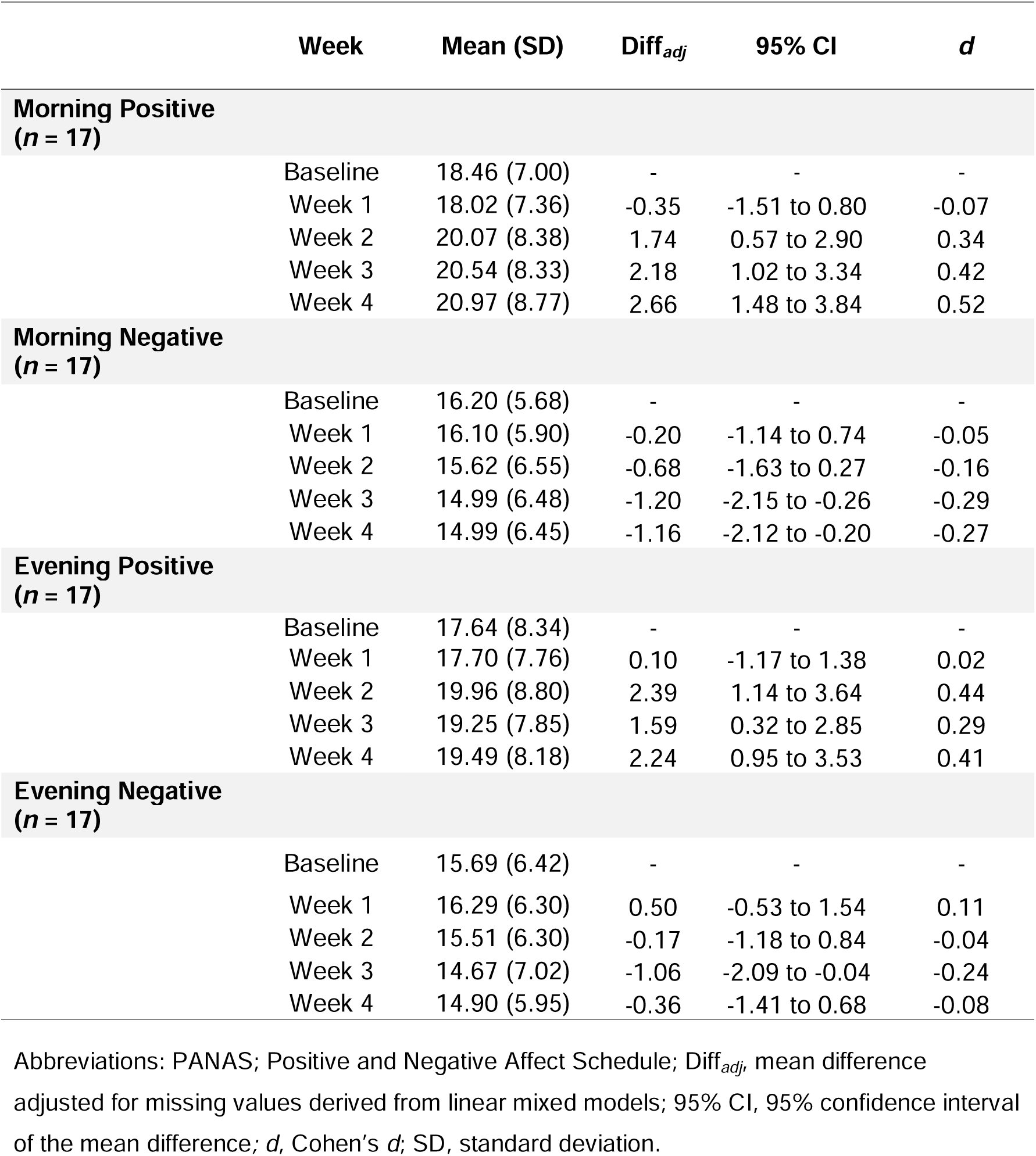
Positive and negative affect (PANAS) by study week.

|  | Week | Mean (SD) | Diff<sub>adj</sub> | 95% CI | *d* |
| --- | --- | --- | --- | --- | --- |
| **Morning Positive**<br>**(*n* = 17)** |  |  |  |  |  |
|  | Baseline | 18.46 (7.00) | - | - | - |
|  | Week 1 | 18.02 (7.36) | -0.35 | -1.51 to 0.80 | -0.07 |
|  | Week 2 | 20.07 (8.38) | 1.74 | 0.57 to 2.90 | 0.34 |
|  | Week 3 | 20.54 (8.33) | 2.18 | 1.02 to 3.34 | 0.42 |
|  | Week 4 | 20.97 (8.77) | 2.66 | 1.48 to 3.84 | 0.52 |
| **Morning Negative**<br>**(*n* = 17)** |  |  |  |  |  |
|  | Baseline | 16.20 (5.68) | - | - | - |
|  | Week 1 | 16.10 (5.90) | -0.20 | -1.14 to 0.74 | -0.05 |
|  | Week 2 | 15.62 (6.55) | -0.68 | -1.63 to 0.27 | -0.16 |
|  | Week 3 | 14.99 (6.48) | -1.20 | -2.15 to -0.26 | -0.29 |
|  | Week 4 | 14.99 (6.45) | -1.16 | -2.12 to -0.20 | -0.27 |
| **Evening Positive**<br>**(*n* = 17)** |  |  |  |  |  |
|  | Baseline | 17.64 (8.34) | - | - | - |
|  | Week 1 | 17.70 (7.76) | 0.10 | -1.17 to 1.38 | 0.02 |
|  | Week 2 | 19.96 (8.80) | 2.39 | 1.14 to 3.64 | 0.44 |
|  | Week 3 | 19.25 (7.85) | 1.59 | 0.32 to 2.85 | 0.29 |
|  | Week 4 | 19.49 (8.18) | 2.24 | 0.95 to 3.53 | 0.41 |
| **Evening Negative**<br>**(*n* = 17)** |  |  |  |  |  |
|  | Baseline | 15.69 (6.42) | - | - | - |
|  | Week 1 | 16.29 (6.30) | 0.50 | -0.53 to 1.54 | 0.11 |
|  | Week 2 | 15.51 (6.30) | -0.17 | -1.18 to 0.84 | -0.04 |
|  | Week 3 | 14.67 (7.02) | -1.06 | -2.09 to -0.04 | -0.24 |
|  | Week 4 | 14.90 (5.95) | -0.36 | -1.41 to 0.68 | -0.08 |

Abbreviations: PANAS; Positive and Negative Affect Schedule; Diff<sub>adj</sub>, mean difference adjusted for missing values derived from linear mixed models; 95% CI, 95% confidence interval of the mean difference; *d*, Cohen's *d*; SD, standard deviation.

### 3.6. Sleep Diary and Actigraphy-derived Sleep Continuity

Self-reported TIB reduced during treatment weeks compared to baseline (*d* = −2.52 to −1.71). TST decreased only in week one (*d* = −0.83). SOL (*d* = −0.75 to −1.21) and WASO (*d* = −0.70 to −1.10) decreased across treatment weeks while SE increased (*d* = 1.37 to 1.73). Actigraphy data from two participants were excluded due to non-wear across both periods, resulting in *n* = 15. Among these 15, one participant had missing baseline data but provided SRT data, while another had missing SRT data but provided baseline data. These were both due to device failure. As linear mixed models are robust to missing data, all 15 participants were retained in the analysis (Table 4). There were reductions in TIB (*d* = −0.72 to –1.85), TST (*d* = −0.65 to −1.32), SOL (*d* = −0.64 to −0.78) and WASO (*d* = −0.36 to –3.38, apart from Week 3) relative to baseline. Sleep efficiency increased (*d* = 0.48 to 1.61) relative to baseline. There were no reliable differences in sleep midpoint.

**Table 4:** Low-density-EEG-derived Sleep Variables.

|  | Mean (SD) | Diff<sub>adj</sub> | 95% CI | d |
| --- | --- | --- | --- | --- |
| **TIB (mins)** |  |  |  |  |
| Baseline | 461.18 (72.62) | - | - | - |
| Night 1 | 382.33 (67.19) | -78.86 | -121.35 to -36.36 | -0.81 |
| Night 2 | 363.88 (62.42) | -95.27 | -139.45 to -51.10 | -0.95 |
| Night 3 | 334.31 (77.99) | -124.78 | -168.14 to -81.43 | -1.46 |
| Night 4 | 364.07 (50.68) | -91.43 | -135.59 to -47.26 | -1.02 |
| Night 5 | 357.99 (52.88) | -101.11 | -144.47 to -57.76 | -0.96 |
| Night 6 | 401.44 (69.30) | -60.16 | -106.27 to -14.05 | -0.58 |
| Night 7 | 379.77 (46.62) | -78.00 | -124.12 to -31.89 | -0.75 |
| Night 28 | 406.52 (58.56) | -51.90 | -96.07 to -7.74 | -0.48 |
| **TST (mins)** |  |  |  |  |
| Baseline | 414.88 (70.88) | - | - | - |
| Night 1 | 328.41 (80.16) | -86.47 | -138.86 to -34.09 | -0.72 |
| Night 2 | 327.17 (71.98) | -87.10 | -141.53 to -32.67 | -0.80 |
| Night 3 | 308.83 (81.92) | -102.80 | -157.23 to -48.38 | -1.26 |
| Night 4 | 313.02 (91.14) | -98.64 | -153.06 to -44.22 | -1.00 |
| Night 5 | 330.00 (61.41) | -84.62 | -138.04 to -31.19 | -0.83 |
| Night 6 | 366.27 (82.65) | -52.56 | -109.37 to 4.25 | -0.48 |
| Night 7 | 343.91 (49.92) | -66.43 | -123.25 to -9.61 | -0.69 |
| Night 28 | 365.23 (66.59) | -50.36 | -104.78 to 4.06 | -0.45 |
| **SOL (mins)** |  |  |  |  |
| Baseline | 16.73 (14.07) | - | - | - |
| Night 1 | 15.16 (20.45) | -1.56 | -13.08 to 9.96 | -0.06 |
| Night 2 | 17.71 (21.06) | 0.77 | -11.20 to 12.74 | 0.05 |
| Night 3 | 9.67 (12.91) | -6.06 | -18.03 to 5.90 | -0.47 |
| Night 4 | 15.98 (23.83) | -0.33 | -12.30 to 11.63 | 0.04 |
| Night 5 | 7.94 (7.78) | -8.80 | -20.55 to 2.95 | -0.60 |
| Night 6 | 9.07 (8.72) | -8.19 | -20.68 to 4.30 | -0.67 |
| Night 7 | 11.44 (13.82) | -6.42 | -18.91 to 6.07 | -0.45 |
| Night 28 | 12.18 (11.42) | -3.15 | -15.12 to 8.82 | -0.37 |
| **WASO (mins)** |  |  |  |  |
| Baseline | 26.71 (14.15) | - | - | - |
| Night 1 | 15.23 (16.58) | -11.48 | -27.62 to 4.66 | -0.51 |
| Night 2 | 13.42 (14.12) | -13.05 | -29.76 to 3.66 | -0.70 |
| Night 3 | 10.55 (7.57) | -16.34 | -33.05 to 0.37 | -1.18 |
| Night 4 | 14.27 (23.71) | -12.02 | -28.73 to 4.69 | -0.48 |
| Night 5 | 18.16 (29.15) | -8.40 | -24.81 to 8.01 | -0.23 |
| Night 6 | 16.71 (17.62) | -8.90 | -26.32 to 8.52 | -0.35 |
| Night 7 | 16.24 (16.19) | -10.48 | -27.90 to 6.94 | -0.65 |
| Night 28 | 15.82 (20.21) | -10.74 | -27.45 to 5.97 | -0.45 |

| SE (%) |  |  |  |  |
| --- | --- | --- | --- | --- |
| Baseline | 89.83 (4.52) | - | - | - |
| Night 1 | 88.41 (9.71) | -1.42 | -7.82 to 4.98 | -0.12 |
| Night 2 | 90.43 (6.91) | 0.53 | -6.10 to 7.16 | 0.05 |
| Night 3 | 93.02 (5.39) | 3.03 | -3.60 to 9.66 | 0.64 |
| Night 4 | 89.34 (11.04) | -0.70 | -7.33 to 5.93 | -0.10 |
| Night 5 | 92.26 (8.79) | 2.32 | -4.19 to 8.83 | 0.31 |
| Night 6 | 92.86 (7.38) | 2.70 | -4.22 to 9.61 | 0.41 |
| Night 7 | 91.82 (5.13) | 2.28 | -4.64 to 9.19 | 0.54 |
| Night 28 | 92.65 (6.22) | 2.49 | -4.14 to 9.12 | 0.40 |
| N1 (mins) |  |  |  |  |
| Baseline | 18.71 (12.73) | - | - | - |
| Night 1 | 13.70 (6.19) | -5.00 | -11.46 to 1.46 | -0.37 |
| Night 2 | 12.27 (6.78) | -6.32 | -13.03 to 0.38 | -0.43 |
| Night 3 | 8.92 (7.23) | -9.18 | -15.88 to -2.47 | -0.69 |
| Night 4 | 12.33 (12.64) | -6.28 | -12.99 to 0.42 | -0.36 |
| Night 5 | 9.44 (5.50) | -8.95 | -15.53 to -2.36 | -0.71 |
| Night 6 | 8.84 (3.28) | -8.92 | -15.92 to -1.92 | -0.65 |
| Night 7 | 13.14 (9.94) | -5.28 | -12.28 to 1.72 | -0.56 |
| Night 28 | 10.60 (8.26) | -7.65 | -14.36 to -0.95 | -0.55 |
| N2 (mins) |  |  |  |  |
| Baseline | 151.79 (53.09) | - | - | - |
| Night 1 | 114.94 (55.21) | -36.85 | -69.43 to -4.27 | -0.59 |
| Night 2 | 101.30 (36.86) | -52.33 | -86.21 to -18.46 | -1.20 |
| Night 3 | 109.34 (51.01) | -40.90 | -74.76 to -7.03 | -0.73 |
| Night 4 | 98.02 (47.96) | -48.81 | -82.68 to -14.94 | -0.79 |
| Night 5 | 124.68 (41.87) | -26.26 | -59.51 to 6.98 | -0.46 |
| Night 6 | 130.15 (65.37) | -19.51 | -54.87 to 15.85 | -0.25 |
| Night 7 | 129.50 (38.98) | -23.05 | -58.42 to 12.31 | -0.44 |
| Night 28 | 119.61 (46.38) | -28.80 | -62.67 to 5.06 | -0.64 |
| N3 (mins) |  |  |  |  |
| Baseline | 143.12 (51.06) | - | - | - |
| Night 1 | 125.65 (45.52) | -17.47 | -44.30 to 9.36 | -0.37 |
| Night 2 | 136.53 (58.75) | -5.57 | -33.50 to 22.37 | -0.11 |
| Night 3 | 115.27 (55.50) | -27.72 | -55.65 to 0.21 | -0.68 |
| Night 4 | 125.97 (58.56) | -19.60 | -47.53 to 8.34 | -0.50 |
| Night 5 | 127.91 (55.90) | -16.19 | -43.61 to 11.23 | -0.33 |
| Night 6 | 133.23 (42.79) | -15.20 | -44.37 to 13.97 | -0.43 |
| Night 7 | 124.46 (52.30) | -15.37 | -44.55 to 13.80 | -0.22 |
| Night 28 | 153.30 (49.18) | 6.51 | -21.42 to 34.44 | 0.20 |
| REM (mins) |  |  |  |  |
| Baseline | 101.18 (22.05) | - | - | - |
| Night 1 | 74.00 (32.48) | -27.18 | -48.92 to -5.44 | -0.77 |
| Night 2 | 77.07 (31.62) | -22.94 | -45.52 to -0.36 | -0.53 |
| Night 3 | 75.30 (36.78) | -25.05 | -47.62 to -2.47 | -0.59 |
| Night 4 | 76.70 (32.53) | -24.09 | -46.67 to -1.52 | -0.67 |
| Night 5 | 67.94 (25.93) | -33.40 | -55.56 to -11.23 | -1.17 |
| Night 6 | 94.04 (28.92) | -9.40 | -32.96 to 14.17 | -0.18 |
| Night 7 | 76.81 (24.90) | -22.45 | -46.02 to 1.12 | -0.98 |
| Night 28 | 81.72 (26.56) | -20.72 | -43.30 to 1.86 | -0.60 |
| **REM Latency (mins)** |  |  |  |  |
| Baseline | 94.44 (44.94) | - | - | - |
| Night 1 | 81.59 (35.28) | -12.85 | -42.45 to 16.74 | -0.27 |
| Night 2 | 71.17 (38.38) | -24.77 | -55.50 to 5.97 | -0.71 |
| Night 3 | 62.30 (33.38) | -32.84 | -63.56 to -2.11 | -0.63 |
| Night 4 | 53.30 (22.47) | -41.71 | -72.43 to -10.98 | -0.72 |
| Night 5 | 84.12 (43.34) | -10.23 | -40.39 to 19.94 | -0.16 |
| Night 6 | 85.00 (53.68) | -7.74 | -39.81 to 24.33 | -0.15 |
| Night 7 | 64.69 (34.92) | -26.13 | -58.20 to 5.95 | -0.51 |
| Night 28 | 72.77 (36.66) | -20.60 | -51.33 to 10.13 | -0.52 |
| **N3 Latency (mins)** |  |  |  |  |
| Baseline | 32.22 (20.07) | - | - | - |
| Night 1 | 30.31 (24.08) | -1.91 | -16.78 to 12.95 | -0.06 |
| Night 2 | 30.68 (25.64) | -1.79 | -17.24 to 13.65 | -0.06 |
| Night 3 | 26.60 (20.93) | -4.73 | -20.17 to 10.71 | -0.25 |
| Night 4 | 31.58 (25.88) | -0.55 | -15.99 to 14.89 | -0.01 |
| Night 5 | 24.38 (14.79) | -8.08 | -23.24 to 7.08 | -0.39 |
| Night 6 | 27.99 (19.36) | -4.32 | -20.44 to 11.80 | -0.22 |
| Night 7 | 27.82 (18.55) | -5.29 | -21.41 to 10.84 | -0.25 |
| Night 28 | 24.88 (18.25) | -5.83 | -21.27 to 9.61 | -0.33 |

Abbreviations: TIB, time in bed; TST, total sleep time; SOL, sleep onset latency; WASO, wake after sleep onset; SE, sleep efficiency; $\text{Diff}_{adj}$ , mean difference adjusted for missing values derived from linear mixed models; 95% CI, 95% confidence interval of the mean difference; $d$ , Cohen's $d$ ; SD, standard deviation.

**Table 5:** Relative power spectral density (%) by frequency band for maximal common number of epochs of NREM sleep and REM sleep.

| Power Spectral Density (*n* = 17) | Mean (SD) | Diff<sub>adj</sub> | 95% CI | *d* |
| --- | --- | --- | --- | --- |
| **NREM Sleep** |  |  |  |  |
| **Slow Wave Activity (% 0.5-4.5Hz)** |  |  |  |  |
| Baseline | 81.74 (4.08) | - | - | - |
| Night 1 | 81.38 (4.71) | -0.36 | -3.86 to 3.14 | -0.10 |
| Night 2 | 82.28 (6.55) | 0.50 | -3.07 to 4.07 | 0.13 |
| Night 3 | 83.18 (4.87) | 1.28 | -2.36 to 4.92 | 0.34 |
| Night 4 | 83.70 (4.25) | 1.32 | -2.32 to 4.96 | 0.35 |
| Night 5 | 81.16 (6.44) | -0.61 | -4.19 to 2.96 | -0.16 |
| Night 6 | 80.61 (6.31) | -0.66 | -4.56 to 3.24 | -0.18 |
| Night 7 | 81.39 (5.84) | -0.02 | -3.74 to 3.70 | -0.00 |
| Night 28 | 81.49 (6.11) | -0.63 | -4.34 to 3.09 | -0.17 |
| **Delta (% 0.75-4.5Hz)** |  |  |  |  |
| Baseline | 63.62 (5.66) | - | - | - |
| Night 1 | 63.82 (5.77) | 0.20 | -3.15 to 3.55 | 0.06 |
| Night 2 | 64.72 (6.58) | 1.17 | -2.25 to 4.59 | 0.32 |
| Night 3 | 65.52 (4.89) | 1.94 | -1.55 to 5.42 | 0.54 |
| Night 4 | 64.77 (5.47) | 0.33 | -3.16 to 3.81 | 0.09 |
| Night 5 | 64.23 (6.56) | 0.67 | -2.75 to 4.10 | 0.19 |
| Night 6 | 64.22 (9.65) | 1.10 | -2.64 to 4.83 | 0.30 |
| Night 7 | 64.66 (7.62) | 1.33 | -2.23 to 4.89 | 0.37 |
| Night 28 | 65.05 (6.48) | 1.13 | -2.43 to 4.69 | 0.31 |
| **Theta (% 4.5-8.0Hz)** |  |  |  |  |
| Baseline | 8.46 (2.47) | - | - | - |
| Night 1 | 8.58 (2.59) | 0.12 | -1.44 to 1.68 | 0.07 |
| Night 2 | 8.12 (2.52) | -0.32 | -1.92 to 1.27 | -0.19 |
| Night 3 | 7.55 (1.35) | -0.79 | -2.42 to 0.83 | -0.47 |
| Night 4 | 7.78 (2.21) | -0.65 | -2.27 to 0.98 | -0.39 |
| Night 5 | 8.51 (2.70) | 0.07 | -1.52 to 1.66 | 0.04 |
| Night 6 | 9.07 (3.07) | 0.55 | -1.19 to 2.29 | 0.33 |
| Night 7 | 8.35 (2.03) | -0.17 | -1.83 to 1.48 | -0.10 |
| Night 28 | 8.22 (2.26) | -0.05 | -1.71 to 1.60 | -0.03 |
| **Alpha (% 8.0-12.0Hz)** |  |  |  |  |
| Baseline | 4.99 (1.58) | - | - | - |
| Night 1 | 5.06 (1.29) | 0.07 | -0.87 to 1.01 | 0.07 |
| Night 2 | 4.57 (1.62) | -0.38 | -1.34 to 0.59 | -0.37 |
| Night 3 | 4.62 (1.72) | -0.36 | -1.34 to 0.62 | -0.35 |
| Night 4 | 4.20 (1.27) | -0.50 | -1.48 to 0.48 | -0.49 |
| Night 5 | 5.12 (2.00) | 0.18 | -0.79 to 1.14 | 0.17 |
| Night 6 | 4.98 (1.73) | -0.05 | -1.10 to 1.01 | -0.04 |
| Night 7 | 5.04 (2.13) | -0.01 | -1.01 to 1.00 | -0.01 |
| Night 28 | 4.92 (2.07) | 0.06 | -0.94 to 1.06 | 0.06 |
| **Sigma (% 12.0-15.0Hz)** |  |  |  |  |
| Baseline | 1.91 (1.08) | - | - | - |
| Night 1 | 1.99 (1.32) | 0.08 | -0.41 to 0.56 | 0.15 |
| Night 2 | 1.87 (1.35) | -0.07 | -0.56 to 0.43 | -0.13 |
| Night 3 | 2.00 (1.55) | 0.05 | -0.46 to 0.56 | 0.09 |
| Night 4 | 1.58 (0.69) | -0.06 | -0.57 to 0.45 | -0.11 |
| Night 5 | 2.11 (1.53) | 0.17 | -0.33 to 0.67 | 0.33 |
| Night 6 | 2.25 (1.78) | 0.12 | -0.42 to 0.67 | 0.23 |
| Night 7 | 2.08 (1.24) | 0.04 | -0.48 to 0.56 | 0.07 |
| Night 28 | 1.91 (1.08) | - | - | - |
| **Beta (% , 15.0-32.0Hz)** |  |  |  |  |
| Baseline | 2.90 (0.97) | - | - | - |
| Night 1 | 2.99 (0.98) | 0.09 | -1.10 to 1.28 | 0.07 |
| Night 2 | 3.16 (2.08) | 0.26 | -0.95 to 1.47 | 0.20 |
| Night 3 | 2.65 (0.97) | -0.20 | -1.43 to 1.04 | -0.15 |
| Night 4 | 2.74 (0.87) | -0.10 | -1.34 to 1.13 | -0.08 |
| Night 5 | 3.09 (1.55) | 0.19 | -1.02 to 1.40 | 0.15 |
| Night 6 | 3.08 (1.41) | 0.05 | -1.27 to 1.37 | 0.04 |
| Night 7 | 3.14 (1.99) | 0.17 | -1.09 to 1.43 | 0.13 |
| Night 28 | 3.27 (1.69) | 0.42 | -0.84 to 1.68 | 0.33 |
| **REM Sleep** |  |  |  |  |
| **Theta (% , 8.0-12.0Hz)** |  |  |  |  |
| Baseline | 16.22 (4.02) | - | - | - |
| Night 1 | 16.10 (3.87) | -0.11 | -2.36 to 2.13 | -0.05 |
| Night 2 | 14.43 (3.33) | -1.94 | -4.23 to 0.36 | -0.80 |
| Night 3 | 14.34 (2.72) | -2.08 | -4.42 to 0.26 | -0.86 |
| Night 4 | 14.43 (4.04) | -2.29 | -4.67 to 0.10 | -0.95 |
| Night 5 | 15.07 (3.32) | -1.30 | -3.60 to 0.99 | -0.54 |
| Night 6 | 16.36 (3.86) | -0.20 | -2.70 to 2.31 | -0.08 |
| Night 7 | 16.13 (4.00) | -0.19 | -2.58 to 2.20 | -0.08 |
| Night 28 | 15.92 (3.89) | -0.27 | -2.65 to 2.12 | -0.11 |

Abbreviations: Diff<sub>adj</sub>, mean difference adjusted for missing values derived from linear mixed models; 95% CI, 95% confidence interval of the mean difference; *d*, Cohen's *d*; SD, standard deviation.

### 3.7. EEG-Derived Sleep Architecture

TIB and TST (except on Night 6) decreased relative to baseline during the first week of SRT (TIB: d = −0.58 to −1.46; TST: d = −0.69 to −1.26). There were no reliable differences in SE, WASO, or SOL. There were reductions in the amount of N1 on nights 3, 5, and 6 (d = −0.69, −0.71, and −0.65 respectively), and N2 on nights 1, 2, 3, and 4 (d = −0.59, −1.20, −0.73, −0.79 respectively), and REM sleep on all nights except night 6 and 7 (d = −0.18 to −1.17). REM latency decreased on nights 3 and 4 (d = −0.63 and −0.72 respectively). There were no reliable differences in N3 sleep, or N3 sleep latency. On night 28, there was reduced TIB (d = −0.48) and N1 sleep (d = −0.55), relative to baseline. Relative values of sleep stages are reported in the supplement (Table S3).

### 3.8. Power Spectral Analysis

The maximum number of common, artefact-free NREM (N2+N3) epochs across all participants and nights was 162 (1 hour 21 minutes). NREM sleep composition (proportion of N2 and N3) of the trimmed hypnograms is reported for each night in the supplement (Table S4). Excluding one night where a participant had a high artefact percentage during REM sleep, the maximum number of common artefact free epochs was 32 (16 minutes). There were no reliable differences in relative PSD in any frequency band during NREM or in the theta range during REM sleep (Table 6, Figure 5). There were no reliable changes in spindle, or SO count or density (Table 7). The absolute values are reported in the supplement (Table S5).

**Figure 5:**
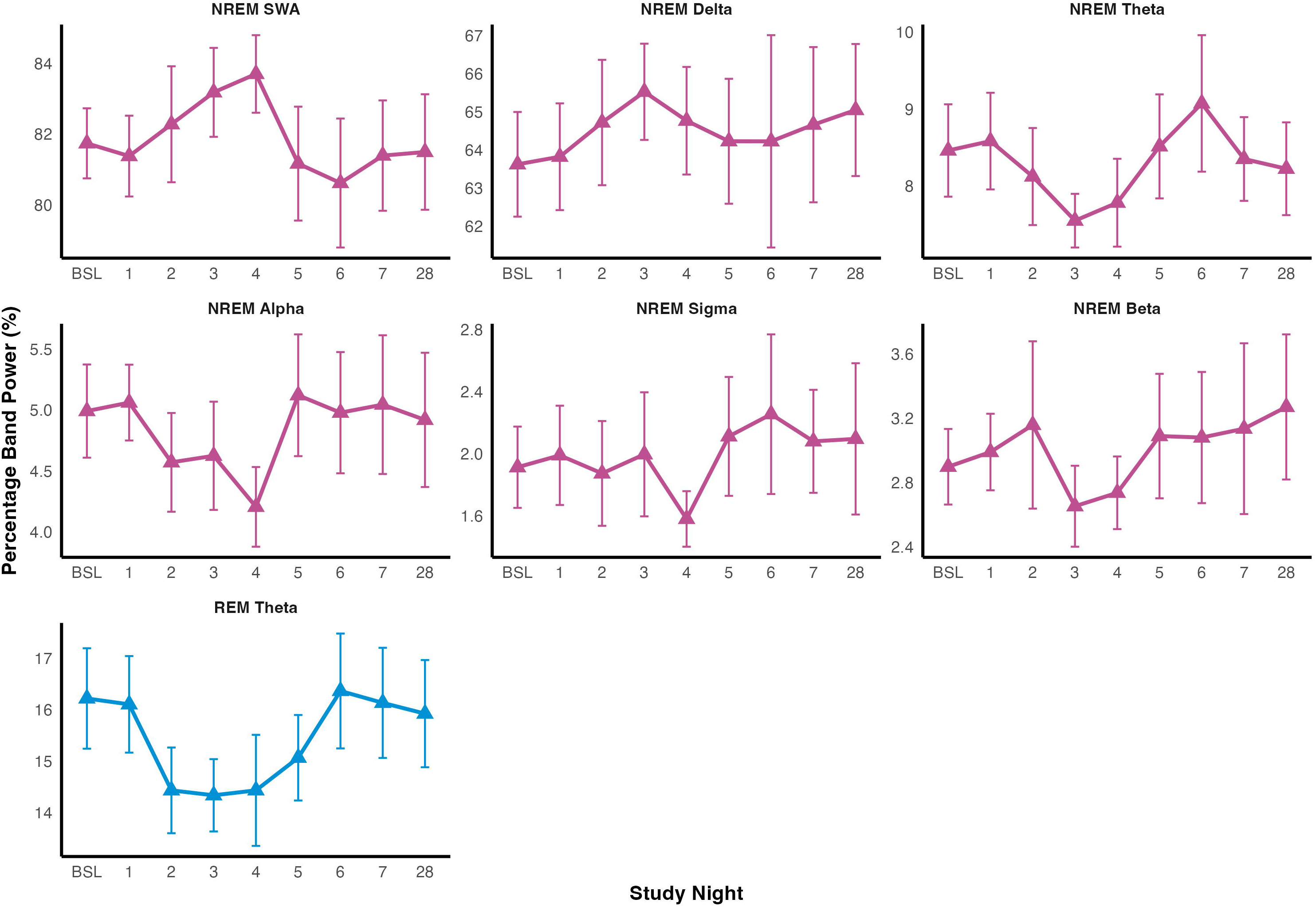
Relative Power Spectral Density (%) in frequency bands in NREM and REM epochs during the first week of SRT. BSL = baseline, Alpha (8.0-12.0Hz), beta (15.0-32.0Hz), theta (4.5-8.0Hz), sigma (12.0-15Hz), delta (0.75-4.5Hz), and slow-wave activity (SWA, 0.5-4.5Hz).

**Table 6:** Spindle and Slow Oscillation Counts and Densities During SRT.

| Timepoint | Mean (SD) | Diff<sub>adj</sub> | 95% CI | *d* |
| --- | --- | --- | --- | --- |
| **Spindle Count (*n* = 17)** |  |  |  |  |
| Baseline | 265.35 (84.49) | - | - | - |
| Night 1 | 250.94 (88.71) | -14.41 | -48.99 to 20.16 | -0.40 |
| Night 2 | 252.21 (102.36) | -16.00 | -52.82 to 20.81 | -0.44 |
| Night 3 | 257.73 (104.94) | -8.25 | -44.28 to 27.79 | -0.23 |
| Night 4 | 249.57 (88.85) | -1.69 | -38.50 to 35.12 | -0.05 |
| Night 5 | 256.38 (95.88) | -7.46 | -42.82 to 27.89 | -0.21 |
| Night 6 | 224.58 (91.08) | -28.84 | -67.48 to 9.79 | -0.80 |
| Night 7 | 252.83 (117.02) | -23.13 | -61.77 to 15.50 | -0.64 |
| Night 28 | 249.71 (99.22) | -10.68 | -47.46 to 26.11 | -0.30 |
| **Spindle Density (*n* = 17)** |  |  |  |  |
| Baseline | 1.64 (0.52) | - | - | - |
| Night 1 | 1.55 (0.55) | -0.09 | -0.30 to 0.12 | -0.40 |
| Night 2 | 1.56 (0.63) | -0.10 | -0.33 to 0.13 | -0.44 |
| Night 3 | 1.59 (0.65) | -0.05 | -0.27 to 0.17 | -0.23 |
| Night 4 | 1.54 (0.55) | -0.01 | -0.24 to 0.22 | -0.05 |
| Night 5 | 1.58 (0.59) | -0.05 | -0.26 to 0.17 | -0.21 |
| Night 6 | 1.39 (0.56) | -0.18 | -0.42 to 0.06 | -0.80 |
| Night 7 | 1.56 (0.72) | -0.14 | -0.38 to 0.10 | -0.64 |
| Night 28 | 1.54 (0.61) | -0.07 | -0.29 to 0.16 | -0.30 |
| **Slow Oscillation Count (*n* = 17)** |  |  |  |  |
| Baseline | 363.71 (89.99) | - | - | - |
| Night 1 | 345.97 (78.62) | -17.74 | -69.68 to 34.20 | -0.33 |
| Night 2 | 361.71 (111.61) | 9.45 | -45.80 to 64.70 | 0.17 |
| Night 3 | 355.30 (82.95) | -8.72 | -62.80 to 45.37 | -0.16 |
| Night 4 | 345.36 (73.66) | -20.57 | -75.82 to 34.68 | -0.38 |
| Night 5 | 345.28 (96.40) | -18.62 | -71.69 to 34.46 | -0.34 |
| Night 6 | 339.50 (119.59) | -24.66 | -82.64 to 33.32 | -0.45 |
| Night 7 | 344.00 (89.77) | -12.02 | -70.01 to 45.96 | -0.22 |
| Night 28 | 379.64 (108.89) | 6.26 | -48.95 to 61.46 | 0.12 |
| **Slow Oscillation Density (*n* = 17)** |  |  |  |  |
| Baseline | 2.25 (0.56) | - | - | - |
| Night 1 | 2.14 (0.49) | -0.11 | -0.43 to 0.21 | -0.33 |
| Night 2 | 2.23 (0.69) | 0.06 | -0.28 to 0.40 | 0.17 |
| Night 3 | 2.19 (0.51) | -0.05 | -0.39 to 0.28 | -0.16 |
| Night 4 | 2.13 (0.45) | -0.13 | -0.47 to 0.21 | -0.38 |
| Night 5 | 2.13 (0.60) | -0.11 | -0.44 to 0.21 | -0.34 |
| Night 6 | 2.10 (0.74) | -0.15 | -0.51 to 0.21 | -0.45 |
| Night 7 | 2.12 (0.55) | -0.07 | -0.43 to 0.28 | -0.22 |
| Night 28 | 2.34 (0.67) | 0.04 | -0.30 to 0.38 | 0.12 |

Abbreviations: Diff<sub>adj</sub>, mean difference adjusted for missing values derived from linear mixed models; 95% CI, 95% confidence interval of the mean difference; *d*, Cohen's *d*; SD, standard deviation.

## 4. Discussion

The aim of this study was to examine the feasibility and acceptability of at-home low-density EEG in individuals with insomnia and depressive symptoms undergoing SRT. The device was well-tolerated, with high completion rates indicating strong participant adherence and sufficient data quality, supporting its suitability for longitudinal, home-based sleep monitoring. SRT was implemented successfully; all participants attended scheduled sessions and adhered to prescribed sleep windows at rates comparable to previous studies^6^. Furthermore, participants reported improved insomnia severity, depressive symptoms, and positive affect.

### 4.1. Feasibility of Low-density At-home EEG

Participants completed 92% of expected nights, although it was not possible to ascertain whether data loss resulted from non-compliance, device malfunction, or cloud connectivity issues. Minor skin irritation (e.g., redness, itching) was reported by two participants but resolved within 3-4 days. This likely resulted from consistent electrode placement in a small area; future studies might incorporate “rest days” to mitigate skin sensitivity. Notably, 137 of 153 recordings (89.5%) were determined to be visually scorable by the researcher with a low average global artefact percentage (2.68%), confirming that self-applied low-density EEG may yield reliable data.

### 4.2. Sleep Continuity

Across the four-week treatment period, increases in self-reported SE were accompanied by significant reductions in TIB, SOL, and WASO. Diary-based TST reduced in week one only, while actigraphy data indicated a reduction in TST relative to baseline across all treatment weeks, consistent with the literature^7^. Furthermore, the stability of sleep midpoint throughout treatment suggests that SRT consolidated sleep without inducing a shift in sleep timing, as previously reported^42^.

During the initial week of SRT, EEG-derived data showed reductions in TIB and TST. We did not detect any reliable changes in SE, SOL, or WASO, although estimates were in the expected direction and often medium to large in effect size for several nights. It is worth noting that baseline SOL (∼16 mins vs ∼20 mins) and WASO were lower (∼26 mins vs ∼59 mins) and SE was higher (∼90% vs ∼82%) compared to those reported in a meta-analysis of laboratory PSG studies of insomnia which may lead to a potential ceiling effect, limiting the scope for further improvement. This may be attributed to the increased comfort of home-based devices or the inclusion of an adaptation night, both of which may have resulted in a baseline night of relatively high-quality sleep. It may also indicate that insomnia is not always associated with PSG-defined alterations in sleep continuity^44^.

### 4.3. Sleep Architecture and Power Spectral Density

During the first week of SRT, reductions were observed in N1, N2, and REM sleep, with the reduction in N1 sleep only persisting at night 28. In contrast, there were no changes in N3 duration, suggesting relative preservation of N3 sleep during sleep restriction. This is consistent with findings in both insomnia patients and healthy controls in experimental sleep restriction and SRT studies^16,45–47^. N3 amount as a proportion of TST, on the other hand, tends to be increased following sleep restriction^16,48^ although we did not find clear evidence across nights for this (see Table S4).

Despite changes in sleep architecture, no reliable differences were observed in relative PSD across NREM frequency bands, relative theta power during REM sleep, or spindle and slow oscillation measures. This contrasts, for example, with a previous study showing increased relative NREM delta power after week 1 and 4 of SRT^16^ and some experimental sleep restriction studies^46,49,50^. However, several sleep restriction studies in healthy participants, where sleep loss is relatively modest, do not show increased delta power/SWA at the beginning of the night^45,51,52^. Methodological factors may account for this, such as the limited number of NREM sleep epochs available for PSD estimation^52^ and modest sample size, which may have constrained sensitivity. Finally, the prevalence of daytime napping could have influenced the results, although we note that self-reported napping was relatively consistent between baseline and SRT weeks 1 and 2.

### 4.4. Insomnia Severity, Depressive symptoms, and Affect

SRT was associated with reductions in insomnia severity and depressive symptoms, consistent with findings from previous work^5–7,9^. Small improvements in daily positive and negative affect further support these findings in line with other studies^10^.

Interestingly, despite average reduction in TST of approximately 83 minutes per night during the first week of SRT participants showed no appreciable decline in positive or negative affect. This contrasts with findings in experimental studies in non-patient samples^53^ suggesting that SRT is suitable for clinical populations experiencing comorbid insomnia and at least mild-to-moderate depressive symptoms.

### 4.5. Strengths and Limitations

Our study shows that participants can successfully self-apply a low-density EEG device at home and record sleep over several consecutive nights. Utilising this novel methodology, we report the first detailed examination of consecutive nights of sleep during the first week of SRT at home. Despite the benefits of home-based sleep monitoring, this approach introduced uncontrolled variability, as consistency of electrode placement within and between individuals was not controlled. Additionally, technical limitations of the EEG device may account for the lack of differences seen in some sleep continuity measures. For instance, “lights-on” and “lights-off” times were inferred from a combination of participant diaries and device light and positional sensors. If participants delayed initiating the recording until they attempted to sleep, rather than 15 minutes prior as instructed - this may have resulted in an underestimation of TIB, and a corresponding inflation of SE. Future studies may benefit from incorporating a behavioural-marker procedure (e.g., perform 5 back and forth eye movements when attempting to initiate sleep) to more accurately determine lights-off timing.

Interpretation may also be limited by the fact that baseline relative N3 sleep was high (M = 31.40%, SD = 11.10%) in comparison to values typically reported in scoring guidelines^37^, healthy individuals, insomnia samples, or individuals with depression^44,54^. This may be due to the use of a single frontal electrode which may overestimate slow-wave amplitude^55^ and artificially inflate N3 when applying the 75µv AASM threshold^56^. This may also explain the lack of changes in spindle metrics; as sleep spindles reach their peak power and density when recorded over central channels. Moreover, given that frontal single-channel devices show poor agreement with PSG for N3 and REM^20,57^, validation studies comparing the HST REM+ with PSG and potential adaptation of AASM scoring for such devices are needed.

Finally, the generalisability of study findings is limited by the demographic profile of the sample, which was highly educated (approximately 59% holding postgraduate qualifications). Results may not be generalisable to diverse insomnia populations. Moreover, whilst we interpreted reliable effects based on 95% CIs - consistent with standard reporting practices for pilot studies, this approach may have de-emphasised certain findings. Therefore, such outcomes warrant formal hypothesis testing in sufficiently powered prospective trials.

### 4.6. Conclusion

These preliminary findings demonstrate the feasibility and acceptability of self-applied, low-density EEG during treatment and highlight the promise of wearable technology for capturing longitudinal, ecologically valid sleep data. They also provide initial mechanistic support for the potential of SRT to improve sleep, affect, and depressive symptoms in individuals with insomnia and depression and lays the groundwork for future controlled studies of SRT to better understand treatment mechanisms.

## Data Availability

All data produced in the present study are available upon reasonable request to the authors.

https://github.com/emstanyer/The-RESTORE-Study

## Acknowledgements

The authors would like to thank Phoebe Homer for assistance with manual scoring of the actigraphy records, and Lucy Jobbins for blinding of the EEG records.

## Author Contributions

E.C.S. was the principal investigator, and conceptualised, designed and coordinated the study, carried out recruitment and statistical analysis and drafted the manuscript. S.D.K., C.A.E., and R.S. contributed to clinical supervision, research conceptualisation and design, and writing of the manuscript. M.L.R. assisted with the ethical approval process, recruitment, and data collection. S.I.R.P. and S.D. conducted the power spectral and spindle analysis. L.T. contributed to the writing of the manuscript. All authors reviewed and approved the final manuscript.

## Pre-registration

This study was pre-registered on Open Science Framework (https://osf.io/3meuz)

## Data analysis

The data analysis scripts used in this project are available on GitHub (https://github.com/emstanyer/The-RESTORE-Study).

