## Supplementary Materials for "Insights from nine nights of self-applied, low-density sleep EEG during sleep restriction therapy for insomnia: a proof-of-concept evaluation"

**Supplementary Table 1: Titration of the prescribed TIB during SRT across study weeks**

|  | **Prescribed sleep window** | | |
| --- | --- | --- | --- |
| **Week** | **Increased (+15 mins) (*n)*** | **Maintained (*n)*** | **Decreased (-15 mins) (*n)*** |
| 1-2 | 13 | 2 | 2 |
| 2-3 | 10 | 7 | 0 |
| 3-4 | 10 | 5 | 2 |

**Supplementary Table 2: Self-reported naps, nap duration, and nap start time**

| **Week** | ***n* (%) Nappers*** | **Mean Naps/Napper** | **Mean Start Time (SD)** | **Mean Length (SD)** |
| --- | --- | --- | --- | --- |
| **Baseline** |  |  |  |  |
| Week 1 | 2 (11.8%) | 2.50 | 14:12 (03:25) | 59.0 (26.1) |
| Week 2 | 6 (35.3%) | 1.33 | 14:35 (03:07) | 34.8 (19.2) |
| **SRT** |  |  |  |  |
| Week 1 | 4 (23.5%) | 1.25 | 14:29 (04:42) | 60.8 (35.2) |
| Week 2 | 4 (23.5%) | 1.25 | 16:58 (04:58) | 42.6 (26.8) |
| Week 3 | 2 (11.8%) | 2.50 | 16:40 (04:17) | 44.4 (17.4) |
| Week 4 | 0 (0.0%) | 0.00 | - | 0.0 (0.0) |

SRT = Sleep Restriction Therapy, † = circular mean, * = this number includes those who reported napping at least once.

**Supplementary Table 3: Low-density-EEG-derived Sleep Stages Absolute Values**

| **EEG % (*n* = 17)** |  | **Mean (SD)** | **Diff_adj_** | **95% CI** | ***d*** |
| --- | --- | --- | --- | --- | --- |
| **N1 (%)** |  |  |  |  |  |
|  | Baseline | 3.89 (2.32) | - | - | - |
|  | Night 1 | 3.62 (1.65) | -0.27 | -1.83 to 1.29 | -0.11 |
|  | Night 2 | 3.30 (1.73) | -0.59 | -2.21 to 1.04 | -0.21 |
|  | Night 3 | 2.55 (1.93) | -1.24 | -2.86 to 0.38 | -0.49 |
|  | Night 4 | 3.37 (3.57) | -0.54 | -2.17 to 1.08 | -0.14 |
|  | Night 5 | 2.61 (1.41) | -1.23 | -2.82 to 0.37 | -0.58 |
|  | Night 6 | 2.19 (0.70) | -1.46 | -3.15 to 0.24 | -0.62 |
|  | Night 7 | 3.53 (2.92) | -0.32 | -2.01 to 1.38 | -0.16 |
|  | Night 28 | 2.56 (1.78) | -1.24 | -2.87 to 0.38 | -0.58 |
| **N2 (%)** |  |  |  |  |  |
|  | Baseline | 32.26 (9.66) | - | - | - |
|  | Night 1 | 29.40 (10.10) | -2.87 | -10.14 to 4.41 | -0.35 |
|  | Night 2 | 27.72 (9.11) | -5.19 | -12.75 to 2.38 | -0.82 |
|  | Night 3 | 32.22 (12.76) | -0.08 | -7.64 to 7.49 | -0.00 |
|  | Night 4 | 26.52 (12.10) | -5.05 | -12.61 to 2.52 | -0.38 |
|  | Night 5 | 35.00 (11.85) | 2.77 | -4.65 to 10.20 | 0.31 |
|  | Night 6 | 31.46 (13.24) | -0.17 | -8.07 to 7.73 | 0.07 |
|  | Night 7 | 34.56 (10.68) | 1.87 | -6.03 to 9.77 | 0.17 |
|  | Night 28 | 29.15 (9.76) | -2.43 | -10.00 to 5.13 | -0.40 |
| **N3 (%)** |  |  |  |  |  |
|  | Baseline | 31.40 (11.10) | - | - | - |
|  | Night 1 | 33.07 (11.40) | 1.68 | -4.19 to 7.54 | 0.19 |
|  | Night 2 | 36.80 (13.01) | 5.54 | -0.56 to 11.65 | 0.66 |
|  | Night 3 | 35.32 (14.24) | 3.47 | -2.64 to 9.57 | 0.40 |
|  | Night 4 | 34.14 (14.26) | 1.68 | -4.42 to 7.78 | 0.18 |
|  | Night 5 | 35.60 (14.40) | 3.79 | -2.20 to 9.78 | 0.36 |
|  | Night 6 | 33.80 (11.22) | 0.96 | -5.41 to 7.34 | 0.04 |
|  | Night 7 | 32.26 (11.26) | 1.40 | -4.98 to 7.77 | 0.16 |
|  | Night 28 | 37.86 (10.95) | 5.30 | -0.80 to 11.40 | 0.84 |
| **REM (%)** |  |  |  |  |  |
|  | Baseline | 22.24 (4.80) | - | - | - |
|  | Night 1 | 19.44 (8.15) | -2.80 | -8.71 to 3.11 | -0.39 |
|  | Night 2 | 21.44 (9.24) | -0.62 | -6.75 to 5.51 | -0.03 |
|  | Night 3 | 22.94 (10.61) | 0.63 | -5.50 to 6.76 | 0.07 |
|  | Night 4 | 21.14 (9.12) | -1.27 | -7.40 to 4.85 | -0.11 |
|  | Night 5 | 18.87 (6.76) | -3.50 | -9.51 to 2.52 | -0.61 |
|  | Night 6 | 23.18 (5.33) | 0.35 | -6.04 to 6.75 | 0.22 |
|  | Night 7 | 20.18 (5.66) | -1.74 | -8.13 to 4.66 | -0.26 |
|  | Night 28 | 20.11 (6.11) | -2.58 | -8.71 to 3.54 | -0.34 |
| 95% CI, 95% confidence interval; Diff*adj*, mean difference adjusted via LMM; *d*, Cohen’s *d*; SD, standard deviation. | | | | | |

**Supplementary Table 4: Composition of Trimmed NREM Epochs (N2 vs N3)**

| **Condition** | **N2 (%)** | **N3 (%)** |
| --- | --- | --- |
| Baseline | 36.1 | 63.9 |
| Night 1 | 34.7 | 65.3 |
| Night 2 | 34.1 | 65.9 |
| Night 3 | 36.3 | 63.7 |
| Night 4 | 38.4 | 61.6 |
| Night 5 | 39.0 | 61.0 |
| Night 6 | 39.2 | 60.8 |
| Night 7 | 32.3 | 67.7 |
| Night 28 | 32.7 | 67.3 |

**Supplementary Table 5: Absolute Power Spectral Density (µv^2^) by Frequency Band for NREM and REM Sleep**

| **Power Spectral Density (*n* = 17)** | | **Mean (SD)** | **Diff_adj_** | | **95% CI** | | ***d*** | |
| --- | --- | --- | --- | --- | --- | --- | --- | --- |
| **NREM Sleep** | | | | | | | | |
| **Slow Wave Activity (µV^2^, 0.5-4.5Hz)** | |  |  | |  | |  | |
| Baseline | | 227.70 (121.51) | - | | - | | - | |
| Night 1 | | 255.49 (189.35) | 27.79 | | -76.61 to 132.19 | | 0.25 | |
| Night 2 | | 273.35 (183.03) | 42.54 | | -64.12 to 149.20 | | 0.38 | |
| Night 3 | | 254.36 (177.70) | 19.10 | | -89.56 to 127.77 | | 0.17 | |
| Night 4 | | 307.10 (262.30) | 74.76 | | -33.91 to 183.43 | | 0.67 | |
| Night 5 | | 265.86 (241.11) | 35.06 | | -71.60 to 141.72 | | 0.31 | |
| Night 6 | | 242.94 (155.08) | 3.37 | | -113.05 to 119.80 | | 0.03 | |
| Night 7 | | 234.48 (136.11) | 18.07 | | -92.91 to 129.04 | | 0.16 | |
| Night 28 | | 253.71 (134.39) | 1.88 | | -109.06 to 112.82 | | 0.02 | |
| **Delta (µV^2^, 0.75-4.5Hz)** | |  |  | |  | |  | |
| Baseline | | 172.94 (101.84) | - | | - | | - | |
| Night 1 | | 172.26 (108.62) | -0.68 | | -56.68 to 55.31 | | -0.01 | |
| Night 2 | | 201.95 (132.86) | 27.08 | | -30.16 to 84.33 | | 0.45 | |
| Night 3 | | 183.18 (122.65) | 5.54 | | -52.78 to 63.87 | | 0.09 | |
| Night 4 | | 208.21 (147.94) | 28.66 | | -29.66 to 86.99 | | 0.48 | |
| Night 5 | | 192.08 (160.43) | 17.21 | | -40.03 to 74.46 | | 0.29 | |
| Night 6 | | 181.25 (132.49) | 0.77 | | -61.73 to 63.27 | | 0.01 | |
| Night 7 | | 175.87 (112.02) | 12.34 | | -47.23 to 71.90 | | 0.21 | |
| Night 28 | | 193.99 (112.06) | 3.27 | | -56.28 to 62.81 | | 0.05 | |
| **Theta (µV^2^, 4.5-8.0Hz)** | |  |  | |  | |  | |
| Baseline | | 8.97 (3.20) | - | | - | | - | |
| Night 1 | | 9.58 (3.75) | 0.61 | | -2.92 to 4.13 | | 0.16 | |
| Night 2 | | 10.24 (5.32) | 1.32 | | -2.28 to 4.92 | | 0.35 | |
| Night 3 | | 8.69 (4.48) | -0.27 | | -3.93 to 3.40 | | -0.07 | |
| Night 4 | | 10.26 (6.06) | 1.36 | | -2.31 to 5.02 | | 0.36 | |
| Night 5 | | 10.09 (7.61) | 1.17 | | -2.43 to 4.77 | | 0.31 | |
| Night 6 | | 8.92 (3.57) | -0.12 | | -4.05 to 3.81 | | -0.03 | |
| Night 7 | | 11.31 (11.88) | 2.93 | | -0.82 to 6.67 | | 0.77 | |
| Night 28 | | 10.23 (4.12) | 0.79 | | -2.95 to 4.54 | | 0.21 | |
| **Alpha (µV^2^, 8.0-12.0Hz)** | |  |  | |  | |  | |
| Baseline | | 5.00 (2.07) | - | | - | | - | |
| Night 1 | | 5.72 (3.15) | 0.73 | | -1.49 to 2.94 | | 0.30 | |
| Night 2 | | 5.60 (3.18) | 0.65 | | -1.61 to 2.91 | | 0.27 | |
| Night 3 | | 5.07 (3.04) | -0.03 | | -2.33 to 2.28 | | -0.01 | |
| Night 4 | | 5.56 (3.99) | 0.91 | | -1.39 to 3.21 | | 0.38 | |
| Night 5 | | 5.78 (4.25) | 0.83 | | -1.43 to 3.09 | | 0.35 | |
| Night 6 | | 4.70 (2.78) | -0.41 | | -2.88 to 2.06 | | -0.17 | |
| Night 7 | | 6.56 (8.04) | 1.81 | | -0.54 to 4.16 | | 0.76 | |
| Night 28 | | 5.79 (2.25) | 0.56 | | -1.79 to 2.91 | | 0.24 | |
| **Sigma (µV^2^, 12.0-15.0Hz)** | |  |  | |  | |  | |
| Baseline | | 1.88 (1.26) | - | | - | | - | |
| Night 1 | | 2.29 (2.11) | 0.41 | | -0.90 to 1.72 | | 0.29 | |
| Night 2 | | 2.36 (1.83) | 0.46 | | -0.87 to 1.80 | | 0.33 | |
| Night 3 | | 2.01 (1.55) | 0.06 | | -1.30 to 1.42 | | 0.04 | |
| Night 4 | | 2.12 (1.70) | 0.54 | | -0.82 to 1.90 | | 0.38 | |
| Night 5 | | 2.36 (2.20) | 0.46 | | -0.88 to 1.80 | | 0.33 | |
| Night 6 | | 2.27 (2.35) | 0.15 | | -1.31 to 1.61 | | 0.10 | |
| Night 7 | | 3.00 (4.27) | 1.10 | | -0.29 to 2.50 | | 0.78 | |
| Night 28 | | 2.43 (1.78) | 0.40 | | -1.00 to 1.79 | | 0.28 | |
| **Beta (µV^2^, 15.0-32.0Hz)** | |  |  | |  | |  | |
| Baseline | | 3.19 (1.57) | - | | - | | - | |
| Night 1 | | 4.62 (3.70) | 1.43 | | -2.22 to 5.07 | | 0.38 | |
| Night 2 | | 4.74 (3.89) | 1.55 | | -2.09 to 5.20 | | 0.41 | |
| Night 3 | | 3.59 (2.84) | 0.34 | | -3.37 to 4.05 | | 0.09 | |
| Night 4 | | 4.90 (5.39) | 1.88 | | -1.83 to 5.59 | | 0.49 | |
| Night 5 | | 4.61 (6.42) | 1.42 | | -2.23 to 5.06 | | 0.37 | |
| Night 6 | | 3.64 (2.72) | 0.17 | | -3.80 to 4.14 | | 0.05 | |
| Night 7 | | 5.21 (8.68) | 2.15 | | -1.64 to 5.94 | | 0.57 | |
| Night 28 | | 4.88 (4.11) | 1.43 | | -2.35 to 5.22 | | 0.38 | |
| **REM Sleep** | | | | | | | | |
| **Theta (µV^2^, 4.5-8.0Hz)** |  | | |  | |  | |  |
| Baseline | 5.74 (2.45) | | | - | | - | | - |
| Night 1 | 7.09 (3.85) | | | 1.35 | | -2.82 to 5.51 | | 0.30 |
| Night 2 | 5.73 (2.97) | | | 0.06 | | -4.19 to 4.30 | | 0.01 |
| Night 3 | 6.14 (4.93) | | | 0.30 | | -4.03 to 4.62 | | 0.07 |
| Night 4 | 6.81 (4.23) | | | 0.96 | | -3.46 to 5.37 | | 0.21 |
| Night 5 | 6.53 (3.93) | | | 0.86 | | -3.39 to 5.11 | | 0.19 |
| Night 6 | 5.52 (2.16) | | | -0.57 | | -5.21 to 4.06 | | -0.13 |
| Night 7 | 10.03 (14.26) | | | 4.37 | | -0.05 to 8.78 | | 0.98 |
| Night 28 | 5.81 (2.51) | | | -0.08 | | -4.50 to 4.33 | | -0.02 |

**Actigraphy Analysis Procedure**

In brief, actigraphy timestamps were adjusted for Daylight Savings Time, then sleep windows were manually scored using diary entries and marker buttons pressed by participants when getting into and out of bed. Periods in which the participant reported that the watch was removed, as well as visually identified gaps of ≥ three hours, were marked as missing data. Recordings were included in the sleep analysis only if they contained at least four scored sleep periods. Sleep midpoint was exported in minutes from midnight by the software (MotionWare), averaged as a circular, decimal time and then converted to a clock time (HH:MM).

**Adherence Calculations**

Adherence to the prescribed sleep schedule was assessed by comparing prescribed bed and rise times to those recorded in the sleep diary and/or derived from actigraphy. For each day, participants were considered adherent if the reported or actigraphy-derived bedtime was no more than 15 minutes earlier than prescribed, and if the reported or actigraphy-derived risetime was no more than 15 minutes later than the prescribed risetime.

Each day was scored as follows:

- 2 points if both bedtime and risetime were adherent,
- 1 point if only one time was adherent,
- 0 points if neither time was adherent.

**Overall adherence** was calculated using only days with both bedtime and risetime recorded. **Adherence** rates were also computed separately for bedtime and risetime; thus, a day contributed if a relevant time was present, even if the other was missing.

Overall percentage adherence was computed as:

$$\text{Adherence (\%)}=\frac{\text{Sum of daily adherence points}}{\text{Number of days with both bedtime and risetime recorded × 2}}\times100$$

Only participants with ≥14 completed diary days were included in the analysis.

**Supplementary Figure Legends**

**Supplementary Figure 1: Adherence to the prescribed sleep window across SRT treatment weeks**

A) Bars represent median clock time in HH:MM, with the lighter shaded area representing the IQR. Sleep schedule is taken from the time fell asleep to the time woken up derived from the actiwatch. B) Time in bed derived from sleep diary and actigraphy compared to the prescribed time in bed during SRT. Circles represent the prescribed time given during SRT. Squares represent the time in bed derived from actigraphy, whereas triangles represent the time in bed derived from the self-reported sleep diary. Points represent the mean and error bars represent the standard error (SE). BSL = baseline. C) Adherence to the prescribed bed and risetimes across SRT treatment weeks. Difference between actigraphy-derived bed (circles) and risetimes (triangles) and prescribed time during SRT (first panel), and the difference between self-reported sleep diary bed and risetimes and the prescribed times during SRT in minutes (second panel). The dashed horizontal lines represent 15 minutes difference (either earlier or later) than the prescribed time, used in the adherence calculation. Points represent the mean and error bars represent the standard error (SE). D) Adherence to prescribed bed and risetimes across all SRT treatment days. Difference between actigraphy-derived bed (circles) and risetimes (triangles) and prescribed time during SRT (first panel), and the difference between self-reported sleep diary bed and risetimes and the prescribed times during SRT in minutes (second panel). The dashed horizontal lines represent 15 minutes difference (either earlier or later) than the prescribed time, used in the adherence calculation. Points represent the mean and error bars represent the standard error (SE).
