## Supplementary figures and images for "Insights from nine nights of self-applied, low-density sleep EEG during sleep restriction therapy for insomnia: a proof-of-concept evaluation"

### Figure S1

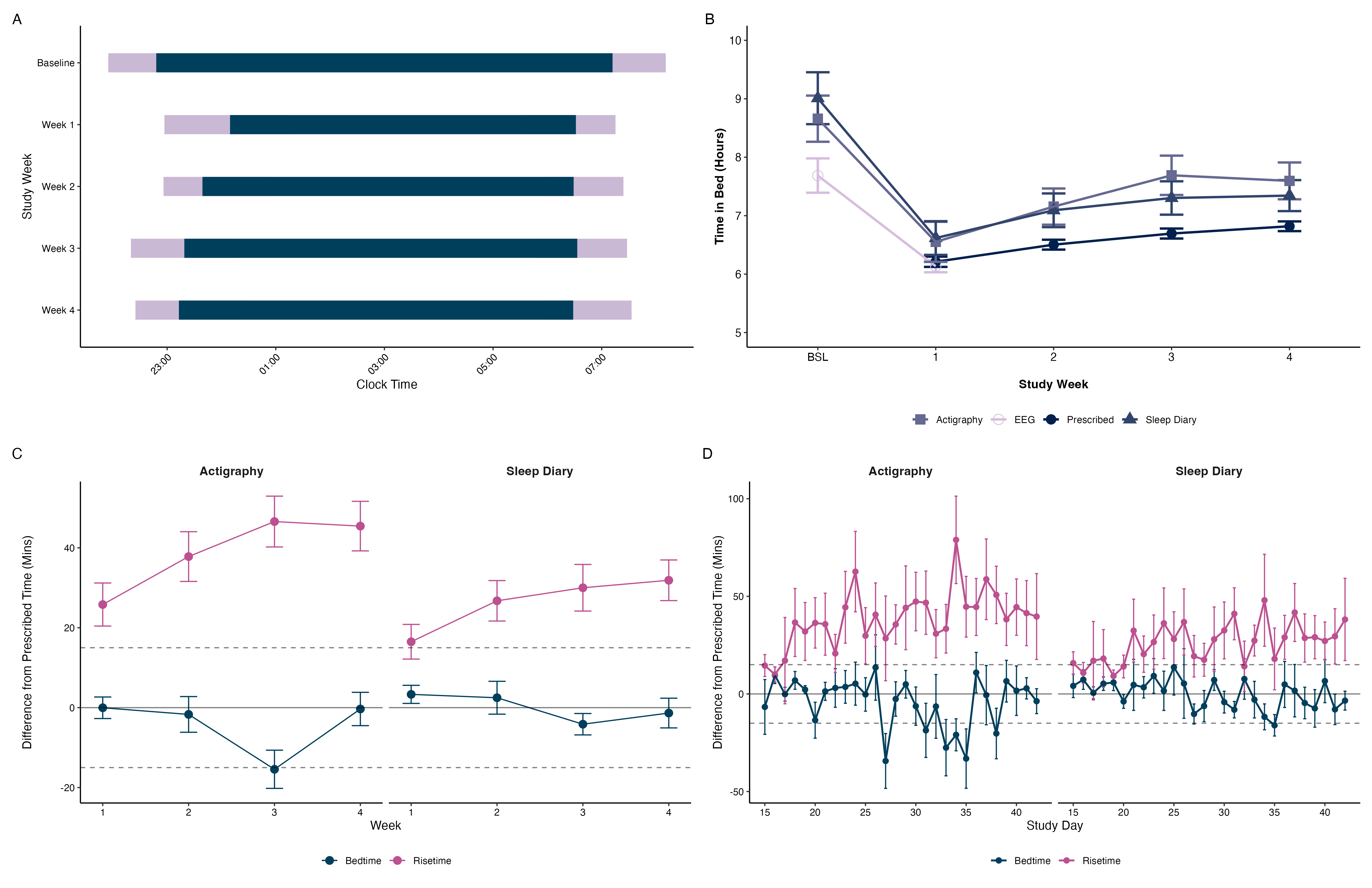
